# Pipeline for Identifying Genetic and Demographic Predictors of Latent Reading Ability: Demonstration in ALSPAC Cohort

**DOI:** 10.1101/2021.08.24.21262573

**Authors:** Hope Sparks Lancaster, Valentin Dinu, Amit Arora, Jing Li, Jeffrey R Gruen

## Abstract

**Purpose:** Reading ability is a complex skill utilizing multiple proficiencies and that develops through interactions between genetic and environmental factors. This study presents an alternative analytic pipeline to identify key genetic and demographic contributors to reading ability.

**Methods:** We analyzed data from the Avon Longitudinal Study of Parents and Children (ALSPAC; N = 3 232) using a multi-step analytical pipeline. To reduce measurement error, we generated a latent reading ability score. We selected single nucleotide polymorphisms (SNPs) based on existing literature and genome-wide association studies (GWAS). We applied elastic net regression to identify informative predictors in two models, a SNP-only model and a SNP-plus demographic, environmental, and behavioral variables model. We compared the SNP-based heritability estimates and R^2^ from the fitted models. We also performed pathway enrichment analysis on the informative SNPs.

**Results:** The traditional GWAS identified one genome-wide significant SNP on chromosome X and produced a moderate heritability estimate of .23 (SE = 0.07). We included 148 SNPs in the elastic net models. The SNP-only model identified 61 informative SNPs (R^2^ = .12), whereas the SNP-plus model identified 96 informative SNPs (R^2^ = .32). The SNP-plus model also showed that several behavioral characteristics positively predicted latent reading ability. Enrichment analysis revealed overrepresentation of several biological pathways among the informative SNPs.

**Conclusions:** This study shows that our analytic pipeline can identify important genetic and demographic predictors of reading ability, providing a powerful alternative to traditional methods and contributing to a deeper understanding of the factors that drive reading development.

---

Efficient and adequate reading depends on several neurocognitive processes and pathways^1^ that support critical component skills. These skills include working memory, phonological awareness,^2^ decoding,^3^ rapid automatized naming (also referred to as fluency)^4^, and language comprehension^3^. Substantial evidence shows that both genetic and non-genetic factors contribute to reading ability^5–7^. Although research on specific genetic variants related to reading continues to evolve, recent studies have made significant progress in this area^8,9^. Behavioral work has also highlighted the influence of various factors on reading development, including demographic characteristics, environmental exposures, and cognitive skills^10^.

Despite this progress, most genetic studies have used traditional approaches with relatively small samples, which limits the identification of relevant genetic predictors. Many studies have also lacked the capacity to integrate genetic and non-genetic data in comprehensive analyses. Three recurring methodological limitations appear in much of the prior work. First, studies often rely on simplified phenotypes that differ across projects. Second, many analyses examine one variable at a time, which limits the ability to detect complex relationships. Third, researchers frequently omit non-genetic factors from analyses that focus on genetic data.

This study aims to demonstrate how alternative analytic methods can address these limitations. By integrating genetic, demographic, environmental, and behavioral data, we seek to identify meaningful associations with reading ability and offer a more comprehensive understanding of its underlying mechanisms.

## Shifting from a Reduced Phenotype to a Multidimensional Phenotype

Many genetic studies use only one or two measures of reading or reading-related tasks to represent reading ability. For example, multiple studies have utilized word reading^11,12^ or single word spelling tasks^13^ as phenotype proxies for reading ability. However, these limited measures fail to capture the full complexity of reading as a multidimensional skill. To address this issue, some researchers have applied multi-trait genetic association methods, such as multi-trait analysis of genome-wide association (MTAG)^8,14^. However, MTAG analyses typically rely on harmonized scores from individual tasks, which may introduce and even amplify measurement error. This discrepancy between observed scores and the underlying true ability could contribute to the lack of replication across genetic studies in different cohorts.

One effective strategy for reducing measurement error involves using latent scores as phenotypes in genetic analysis. A few studies^12,15,16^ have taken this approach by employing principal component analysis (PCA) or composite scores. However, PCA presents several limitations. First, its results may not align with theoretical models and often lack clear interpretability. Second, researchers must decide how many components to retain, which introduces subjectivity. Third, each orthogonal component requires separate interpretation, which can complicate analysis.

Confirmatory factor analysis (CFA) offers a stronger alternative. CFA provides greater control over measurement error, produces interpretable and theoretically grounded scores, and supports replication within and across datasets. Like PCA, CFA allows researchers to compare the fit of multiple models and select the one that best suits the data. In this study, we used CFA to generate a latent reading ability score. This approach enabled us to create a phenotype that is both adaptable and grounded in theory, while also remaining sensitive to the structure of the data.

## Moving from One-At-A-Time to Many-At-Once

Previous studies have established genetic contributions to performance on reading-related quantitative traits, such as nonword reading^9^. Like other complex phenotypes, reading ability reflects the combined influence of many genes. However, most genetic studies have used one-variant-at-a-time statistical approaches, such as genome-wide association studies (GWAS). These methods assess each genetic variant independently and apply strict multiple testing corrections, which often result in data loss—especially in the small sample sizes common in reading genetics research. Because of these stringent thresholds, most studies fail to identify any genetic variants, such as single nucleotide polymorphisms (SNPs), that survive correction. This limitation has slowed progress in uncovering the genetic architecture of reading.

Troung et al.^17^ found only one SNP that passed multiple test correction for rapid automatized naming, a skill that predicts later reading ability. Similarly, Eising et al.^18^ conducted a large meta-GWAS of multiple reading-related traits and identified only one SNP that survived multiple test correction. One-variant-at-a-time approaches also cannot account for interactions or shared variance across genetic markers. Research on developmental dyslexia provides evidence of gene-gene interactions. For instance, variants within READ1 and DCDC2/KIAA0319 have been shown to jointly affect single word reading, nonword reading, spelling, phoneme deletion, and comprehension^19,20^. These findings underscore the importance of analyzing multiple variants together to capture additional predictive value and biological insight.

To address these limitations, researchers can apply statistical models that evaluate many features simultaneously. Machine learning offers one solution, as it can identify informative predictors while accounting for correlations across variables. Elastic net regression, a type of regularized machine learning model, provides an effective tool for this purpose^21,22^. Elastic net models perform feature selection in high-dimensional data and can detect informative genetic variants by leveraging their shared structure and interactions. In this study, we combine a traditional GWAS with an elastic net model to improve the identification of genetic predictors associated with reading ability.

## Incorporating Multiple Data Types

There is evidence for gene-environment interplay for reading development^23,24^. Behavioral research has shown that factors such as biological sex, birth weight, gestational age, maternal education level, and language ability relate to reading outcomes^25^. For example, Mascheretti and colleagues^25^ reported that maternal smoking during pregnancy, birth weight, and socioeconomic status can influence how the DYX1C1 gene affects susceptibility to developmental dyslexia. These findings suggest that reading development arises from a complex interaction between genetic, environmental, and demographic factors.

Despite this complexity, most traditional genetic studies include non-genetic variables only as covariates. This practice limits our ability to explore how genetic and non-genetic factors may jointly contribute to reading development. Research designs and analytic approaches often lack the flexibility to support fully integrated models. As a result, few studies have examined genetic, demographic, and environmental data within the same analysis. To address this gap, we have developed methods that integrate multiple data types using machine learning techniques. In particular, we have applied elastic net models to examine interactions between genetic and environmental factors across various health-related outcomes^26–29^. We have also used these approaches in prior work on reading ability^30^. By applying elastic net modeling to integrate diverse predictors, we aim to advance understanding of how multiple influences contribute to reading ability and to refine hypotheses for future research.

## Study Purpose

This study aimed to demonstrate a pipeline for identifying informative genetic, demographic, and environmental contributors to reading ability. We tested two primary hypotheses. First, we predicted that combining a multidimensional reading phenotype with elastic net models and traditional genetic approaches would identify more informative markers than using traditional methods alone. Second, we hypothesized that the genetic markers identified through this approach would show enrichment in functional biological processes related to brain development. To test these hypotheses, we used data from the Avon Longitudinal Study of Parents and Children (ALSPAC)^31,32^, a well-established population-based cohort. This dataset allowed us to evaluate how our alternative methods can reveal key genetic and non-genetic features associated with reading ability.

## Material and Methods

### Subjects, Recruitment, and DNA Collection

The Avon Longitudinal Study of Parents and Children (ALSPAC) is a population-based birth cohort that has been described extensively in previous research^31,32^. The cohort includes 15 447 pregnancies, which resulted in 15 658 fetuses. Of these, 14 901 children were alive at one year of age. Beginning at age seven, ALSPAC invited participants to annual "Focus" sessions, which collected a wide range of behavioral measures, including assessments of reading and language skills.

For this study, we used data from parent surveys and behavioral assessments related to reading, language, hearing, and cognition. These data were collected during the Focus sessions conducted at ages seven, eight, and nine. A searchable data dictionary and variable search tool, available on the ALSPAC website (http://www.bristol.ac.uk/alspac/researchers/our-data/), provides full details of the available dataset.

After completing quality control and genotype imputation procedures, ALSPAC included genomic data for 8 237 participants. However, not all participants had the necessary behavioral data for this analysis. For our final sample, we included 3 232 participants with both complete genetic and behavioral data.

Additional details about recruitment, inclusion and exclusion criteria, behavioral assessments, demographic characteristics, and DNA collection procedures are provided in Supplementary Methods and in Supplemental Tables S1 and S2.

### Statistical Analysis

Our analytic pipeline included four primary steps: (1) generating a latent reading ability phenotype using confirmatory factor analysis, (2) screening single nucleotide polymorphisms (SNPs) based on prior knowledge and genome-wide association studies (GWAS), (3) identifying informative SNPs using elastic net modeling, and (4) linking informative SNPs to functional biological processes. We also compared the amount of variance explained by traditional GWAS and elastic net models. To estimate this, we used SNP-based heritability for GWAS and R² values for the elastic net models.

We applied this pipeline in two stages. First, we modeled the predictive value of SNPs alone. Then, we included demographic, environmental, and behavioral variables to evaluate the combined effects of genetic and non-genetic features. Commented code for all data analyses is provided in the Supplementary Information.

#### Confirmatory factor analysis

To construct a latent reading ability phenotype, we used confirmatory factor analysis (CFA) with the lavaan package in R^33^. We selected reading-related variables that included single word reading at ages 7 and 9, nonword reading at age 9, spelling at ages 7 and 9, and the Neale Analysis of Reading Ability (NARA)^34^ measures for rate, accuracy, and comprehension. We applied robust modeling procedures to account for missing data. A regression-based equation was then used to compute the latent reading ability score.

#### SNP screening

To constrain the high-dimensionality of the genomic data, we screened SNPs using two methods: (1) review of prior literature and (2) genome-wide association analyses. We examined candidate gene, GWAS, and family-based studies published between 1990 and 2019 that investigated reading ability or reading disability. From this review, we selected 50 SNPs that were reported at least twice to be associated with reading and that were available in the ALSPAC dataset.

We conducted GWAS using PLINK2^35^, analyzing each chromosome separately following methods established in previous work^36^. We specified a generalized linear model with latent reading ability as the outcome variable. We accounted for genetic similarity^*^ by including the top three principal components. From the GWAS results, we selected the top 100 SNPs based on uncorrected p-values across all chromosomes and included any SNPs with a false discovery rate (FDR-BH) of 0.1 or less for the elastic net analysis.

#### SNP-based heritability

To estimate SNP-based heritability, we combined summary statistics across autosomes from the GWAS. We used linkage disequilibrium score regression (LDSC)^37^ to process the summary statistics and calculate heritability estimates. This procedure involved merging alleles and estimating linkage disequilibrium (LD) scores using the 1000 Genomes Project European HapMap3 reference data.

#### Elastic net modeling

We used elastic net regression to identify informative predictors of reading ability. Elastic net models apply regularization to perform variable selection in high-dimensional settings^21,22^. This approach incorporates two penalty terms into the regression loss function. The first term (L1 norm) forces coefficients of less influential predictors to zero, effectively performing feature selection. The second term (L2 norm) allows the model to retain groups of correlated predictors, such as linked SNPs.

We used five-fold cross-validation to determine the optimal tuning parameters for both penalties. During each iteration, we trained the model on four folds and tested it on the fifth, repeating the process until every fold had served as the test set. We repeated this cross-validation ten times to ensure consistent parameter selection. After identifying the best parameters, we refit the final model using the full dataset to estimate the regression coefficients.

We defined informative features as those with absolute beta values greater than 0.019. We entered these features into linear regression models to estimate the variance explained in reading ability. We then compared the resulting models to a null model and to each other. In Model 1, we included only the SNPs selected during the screening stage. In Model 2, we added demographic, environmental, and behavioral variables. These variables included biological sex, race or ethnicity, nonverbal IQ, birth weight, maternal education level, receptive vocabulary at age 8, receptive language ability, and bilingual language status.

#### Enrichment analysis

To examine the biological relevance of the SNPs identified through elastic net modeling, we mapped them to genes using the g:SNPense tool in g:Profiler^38^. We then performed enrichment analysis using the g:GOSt function within g:Profiler. We selected g:Profiler based on recent evaluations that found it to have the most up-to-date and comprehensive repository of biological pathways, drawing from curated sources such as Gene Ontology and KEGG. We submitted the list of informative SNPs to g:GOSt for functional profiling and interpretation of enriched biological processes.

## Results

### Latent Reading Ability Score

We used confirmatory factor analysis (CFA) to create a latent reading ability score by loading all reading-related variables onto a single factor. The base model showed mixed goodness-of-fit statistics (see Supplementary Table S3). We improved the model fit statistically by adding correlated error terms between observed variables, based on both theoretical rationale and modification indices. However, a comparison between the extracted latent scores from the base and improved models revealed no difference in the final scores, *t*(3231) = – 1.203e–15, *p* = 1.

Figure 1 presents standardized path loadings from the base model. All observed variables loaded significantly onto the latent factor, with standardized coefficients ranging from 0.745 to 0.929. The model accounted for a substantial portion of variance in reading ability, with an average explained variance of 0.667 (Supplementary Table S3). We found no negative variance estimates. These results indicate that the single-factor model without correlated errors provided a strong enough fit to generate the latent reading ability score.

**Figure 1.**
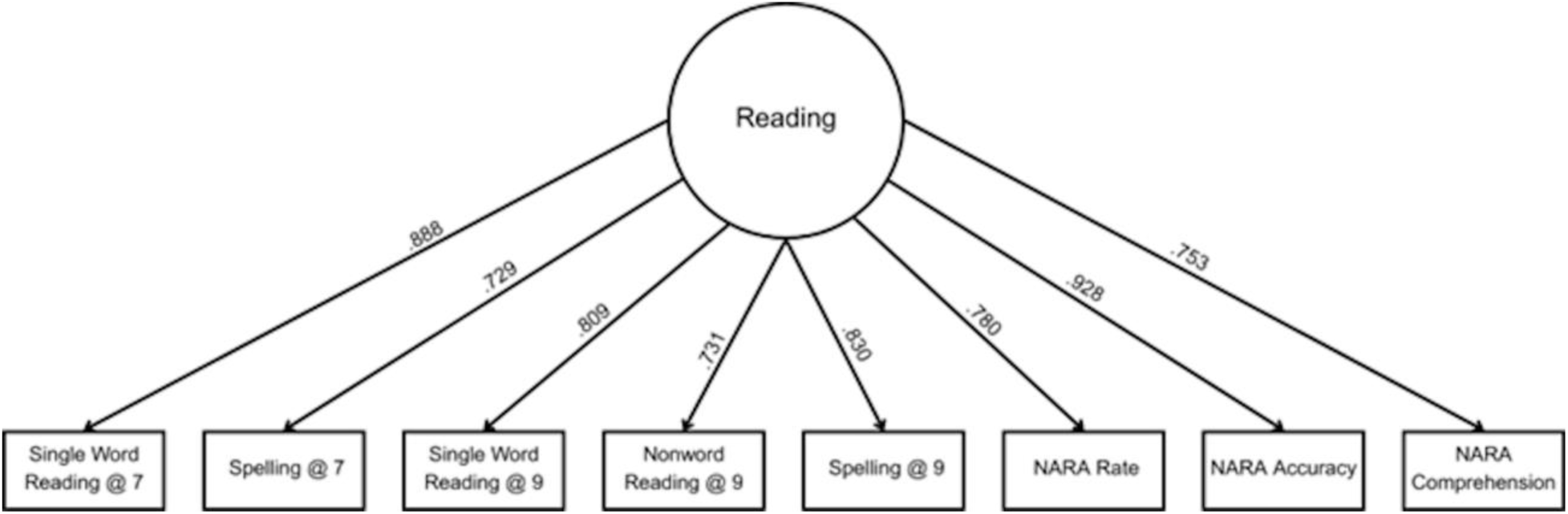
Manifest path values for Latent Reading Ability Base Model for ALSPAC. All paths were significant. NARA = Neale Analysis of Reading Ability (Neale, 1997).

### Genome-Wide Association for SNP Screening

To manage the high dimensionality of genetic data, we used genome-wide association analysis in PLINK2 to screen SNPs before applying multivariate modeling. The analysis used the latent reading ability score as the phenotype. We identified one genome-wide significant SNP after correcting for multiple comparisons: rs181384543 (X:22779030, FDR = 0.007), located within the gene *PTCHD-1AS*.

After combining 101 SNPs identified through GWAS screening with 50 SNPs selected from prior research, we extracted genotype data for 148 SNPs from the ALSPAC dataset (Supplementary Table S4). Three SNPs were unavailable. Using autosomal summary statistics, we estimated a moderate SNP-based heritability of 0.23 (SE = 0.07) for the latent reading ability phenotype.

### Elastic Net Models

We fit two elastic net models to identify informative predictors of reading ability. The first model included only SNP data. The second included SNPs along with demographic, environmental, and behavioral variables. Due to the requirement for complete data in elastic net modeling, our analytic sample decreased (ALSPAC: *n* = 3 647). Informative SNPs are reported in Tables 1 and 2, and model test statistics appear in Supplementary Table S5.

**Table 1.**
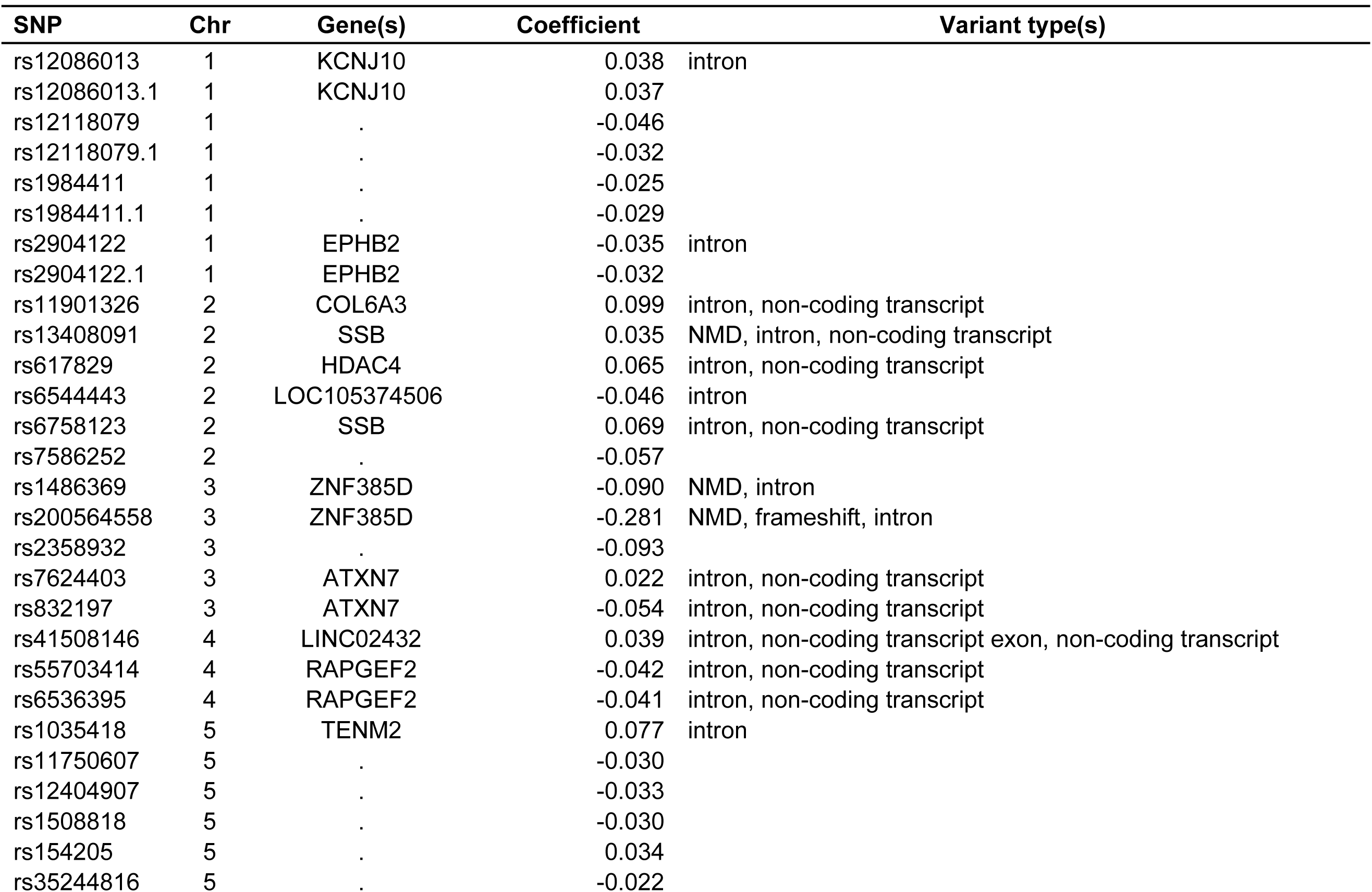
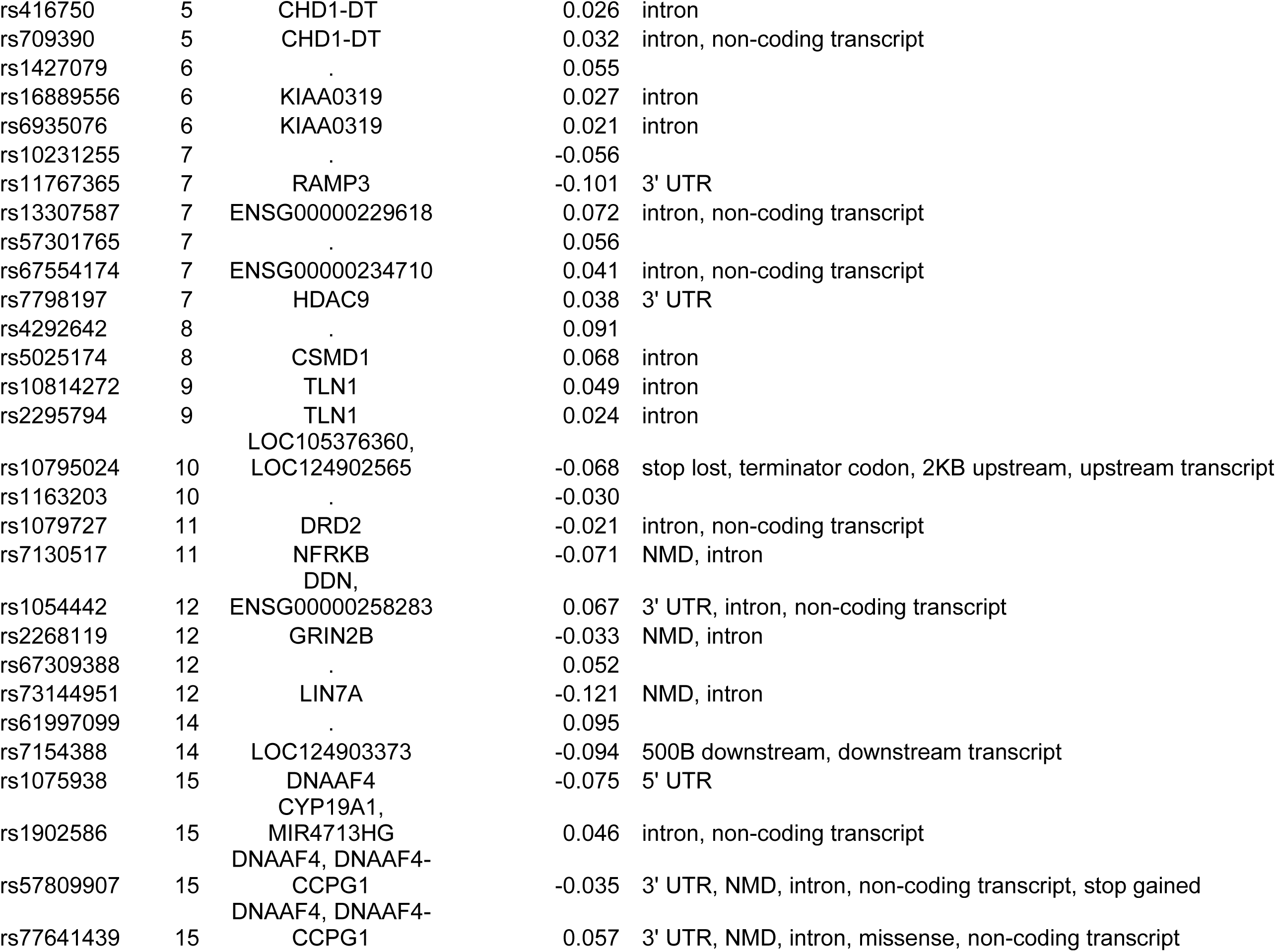
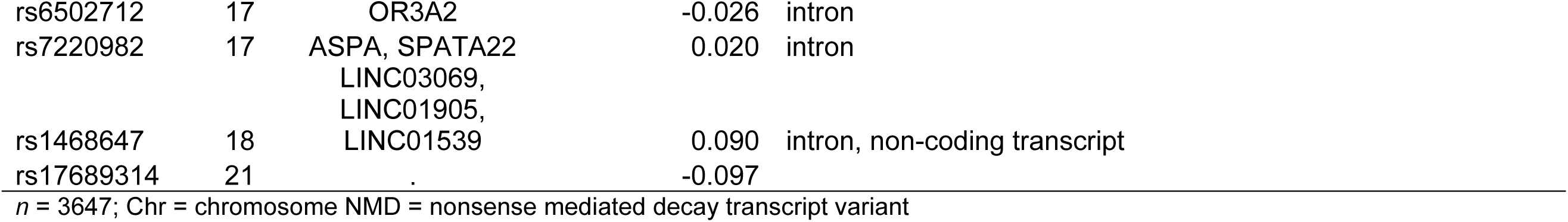
Elastic net results for general reading ability in the ALSPAC - Read SNP only models.

**Table 2.**
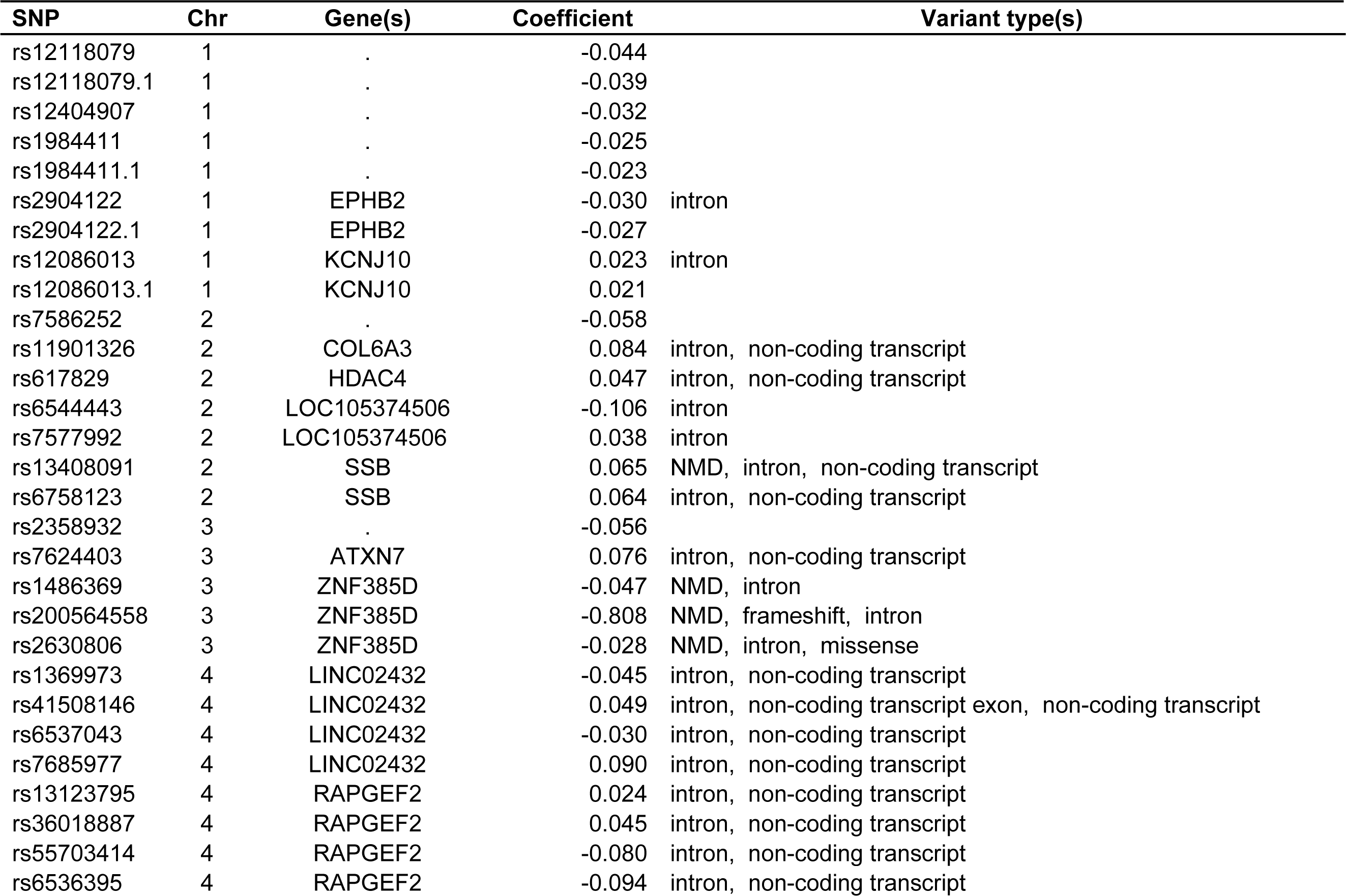
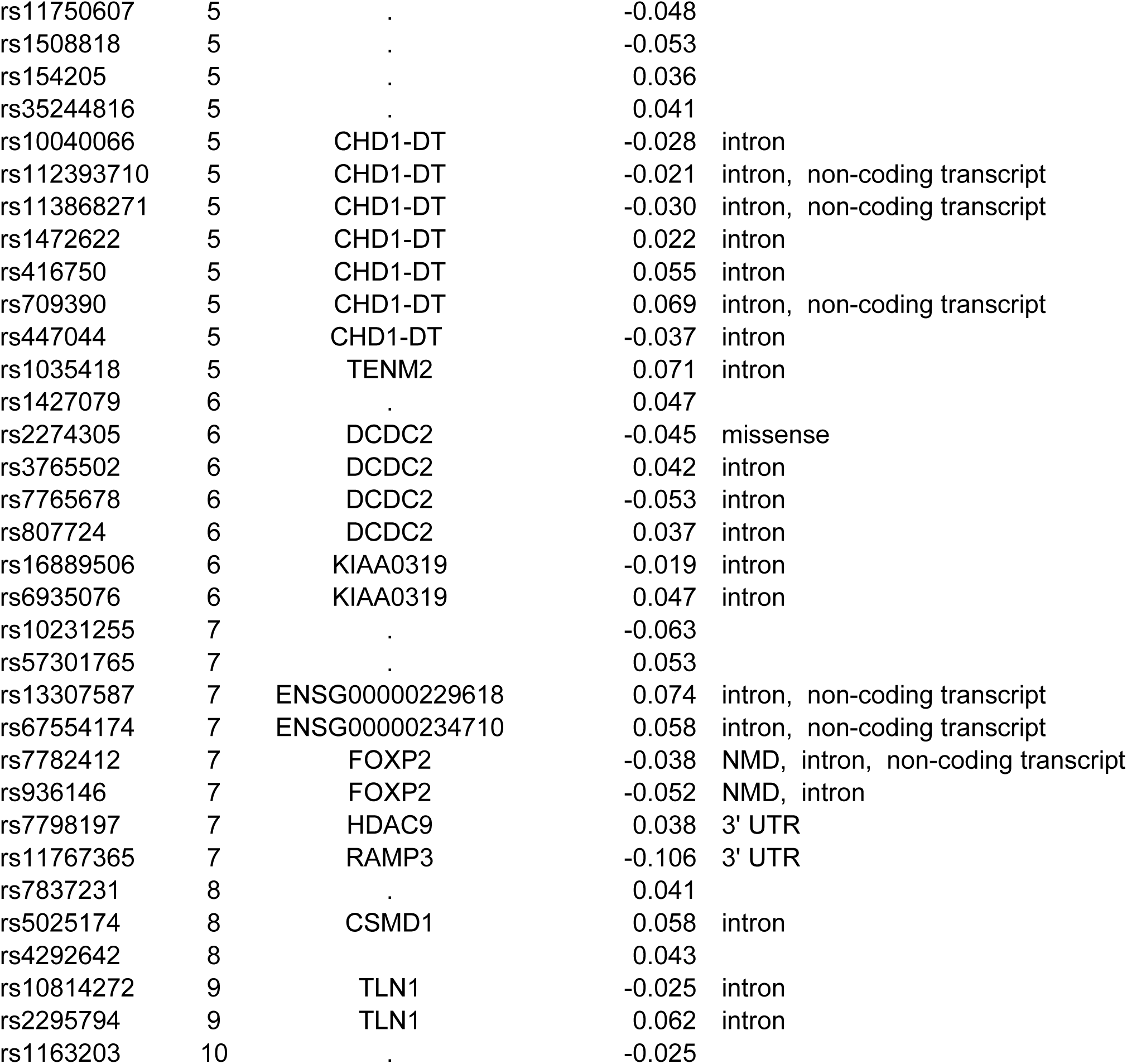
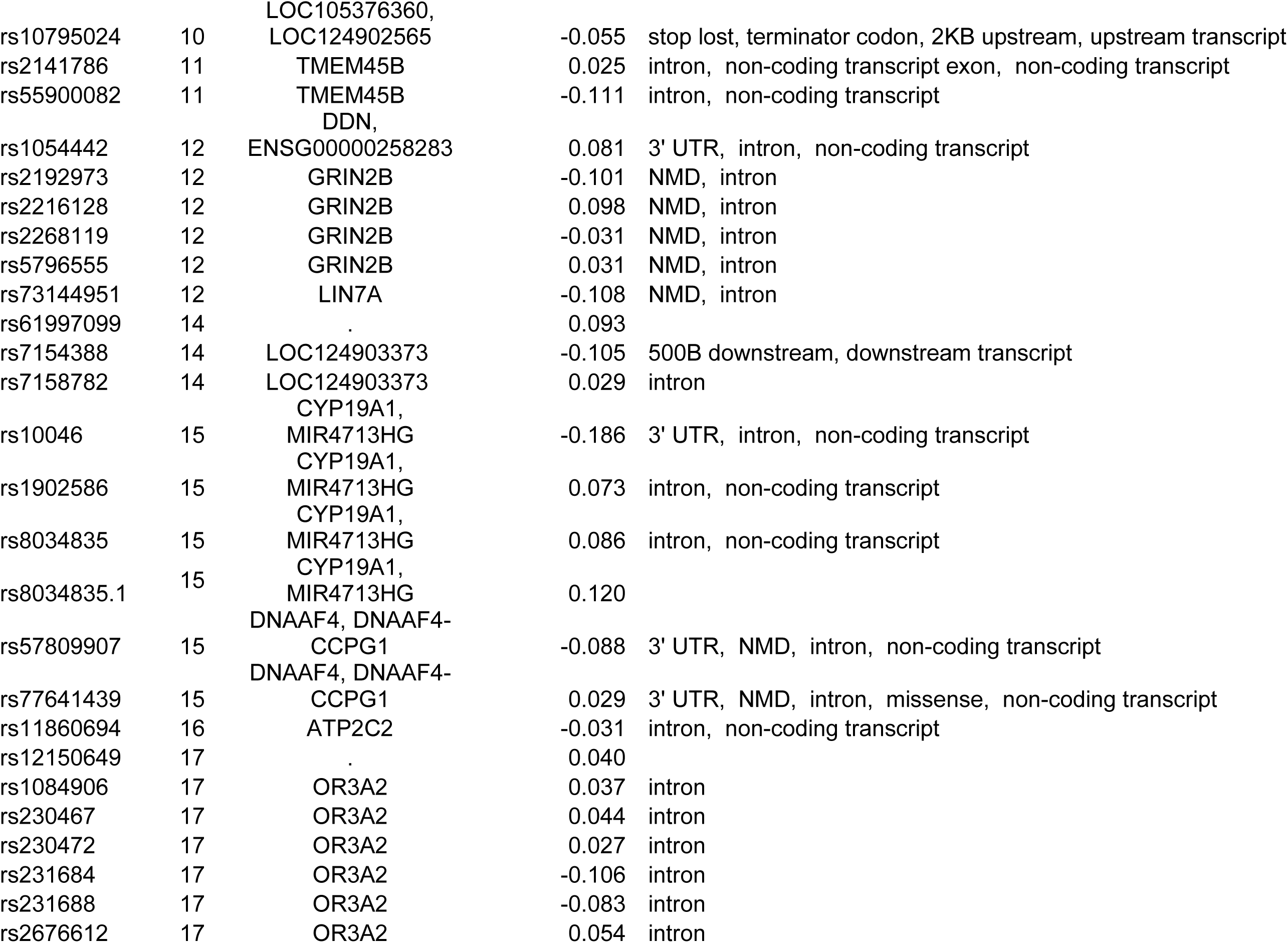
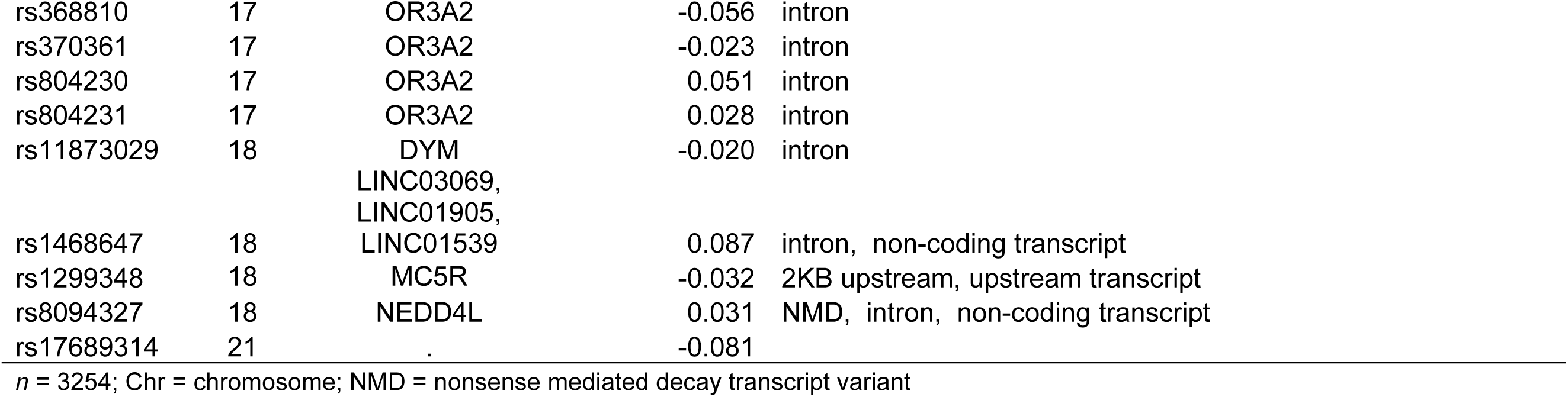
Elastic net results for informative SNPs for general reading ability in the ALSPAC - Read SNP + demographics model.

#### Model 1: SNP-only model

This model identified 62 informative SNPs with absolute beta values greater than 0.019. Thirty-one SNPs had positive coefficients, indicating associations with better reading ability, while 31 had negative coefficients, indicating associations with lower reading ability (Table 1). The positive SNPs mapped to 19 genes and the negative SNPs to 14 genes. Some genes, including *ATCN7* and *DNAAF4*, had both positive and negative associated SNPs.

Most identified SNPs were intronic or non-coding variants. Several SNPs aligned with previous findings in the literature, including variants in *KIAA0319* (chromosome 6), *DRD2* (chromosome 11), *CYP19A1* (chromosome 15), and *DNAAF4* (chromosome 15), supporting replication of prior results from ALSPAC and other cohorts such as the Colorado Reading Disabilities study.

We used linear regression to evaluate model fit and variance explained. The model accounted for 12 percent of variance in latent reading ability, performing significantly better than a null model (Supplementary Table S5; *F*(1, 53) = 10.32, *p* < .0001, Adjusted *R²* = 0.1193). Seven SNP coefficients could not be estimated due to perfect multicollinearity, suggesting redundancy that may allow for model simplification.

#### Model 2: SNP-plus demographic, environmental, and behavioral features model

To assess the combined influence of genetic and non-genetic factors, we added biological sex, race or ethnicity, nonverbal IQ, birth weight, mother’s highest education, receptive vocabulary at age 8, receptive language, and bilingual status to the elastic net model (Table 2). Figure 2 visualizes the relationships between these demographic features and the latent reading ability score.

**Figure 2.**
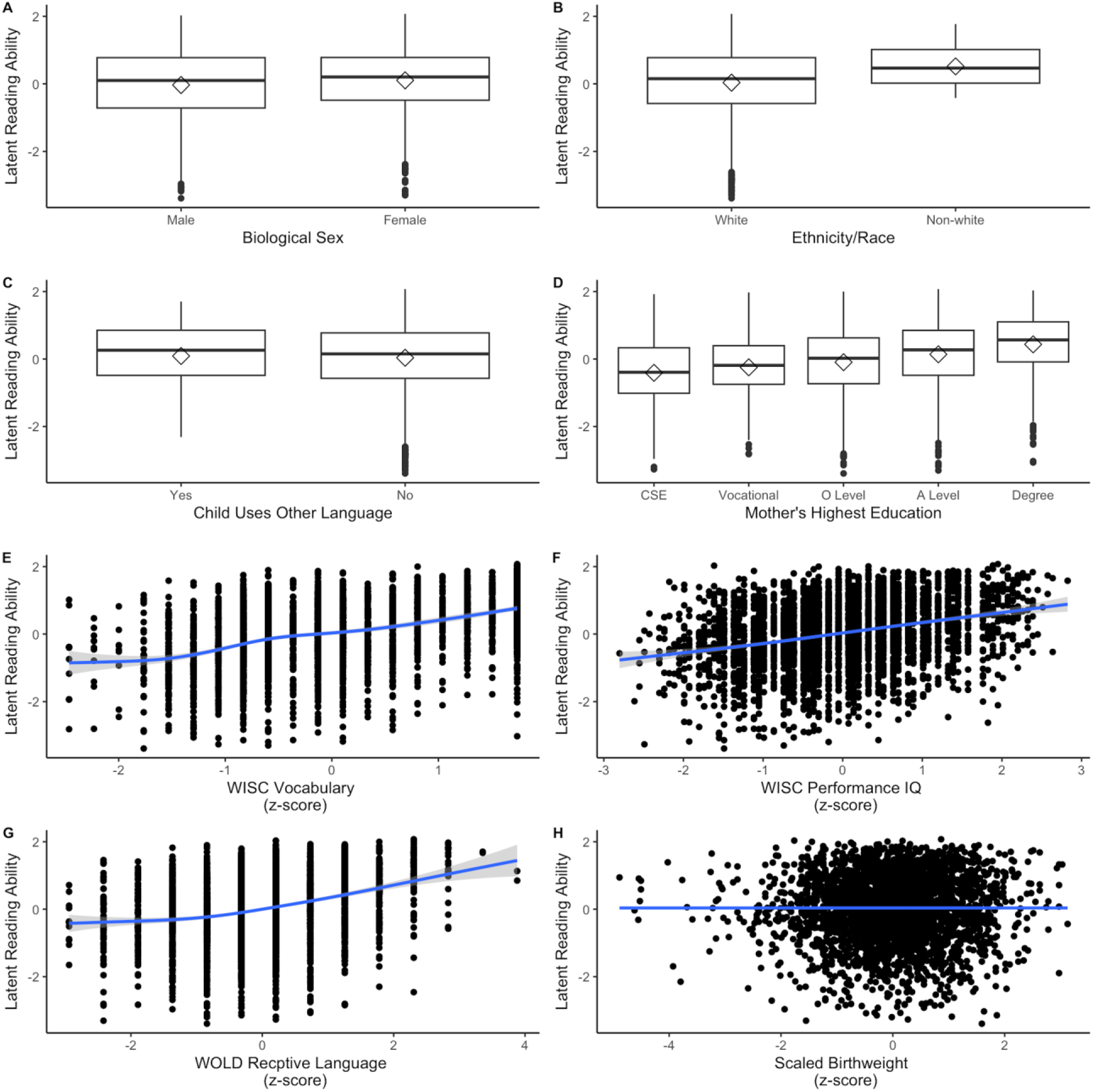
Plots of relationships between demographic variables and latent reading ability score. Plots for the relationships between (A) biological sex, (B) ethnicity / race, (C) use of language other than English, (D) highest level of maternal education, (E) receptive vocabulary, (F) nonverbal intelligence, (G) receptive language, and (H) scaled preferred birthweight in grams and latent reading ability in the ALSPAC cohort.

This model identified 96 informative SNPs, 49 with positive coefficients and 48 with negative coefficients. These SNPs mapped to 31 unique genes across the genome. Twenty-seven of those genes included positive SNPs, and 24 included negative SNPs. Eleven genes included both positive and negative associations, such as *DCDC2*, *KIAA0319*, *GRIN2B*, *CYP19A1*, and *DNAAF4*, all of which have prior associations with dyslexia and reading development. Chromosomes 5 and 17 contributed 24 percent of all informative SNPs, all selected through the GWAS screening stage.

Six demographic features were identified as informative based on beta values exceeding 0.019 in absolute value. These included biological sex, race or ethnicity, nonverbal IQ, receptive language, vocabulary, and mother’s education level. All six features had positive coefficients, indicating that higher values were associated with higher latent reading scores. These findings align with prior studies linking cognitive and environmental variables to reading outcomes.

The model explained 32 percent of the variance in latent reading ability (Supplementary Table S5; *F*(1, 85) = 18.93, *p* < .0001, Adjusted *R²* = 0.3191, *n* = 3254). It outperformed both the null model (*F*(1, 85) = 18.93, *p* < .0001) and the SNP-only model from ALSPAC (*F*(1, 32) = 29.69, *p* < .001). Seventeen SNP coefficients were not estimated due to multicollinearity and were likely redundant with other features in the model.

We also examined correlations between demographic features and latent reading ability. Vocabulary (*r* = .42), nonverbal IQ (*r* = .31), receptive language (*r* = .29), and mother’s education (*r* = .25) showed moderate positive correlations. Biological sex (*r* = .07) and race or ethnicity (*r* = .03) showed weak associations. These weaker correlations suggest that sex and ethnicity may have been selected in the model due to their shared variance with more predictive features.

### Enrichment Analysis

We conducted functional profiling of the informative SNPs using the g:GOSt tool in g:Profiler. The results appear in Table 3. For the SNP-only model, we identified 23 significantly enriched biological pathways (adjusted g:SCS *p* < 0.05). Eight of these pathways were identified as likely drivers, including the postsynaptic membrane (GO:0045211, *p* = .0001) and learning or memory processes (GO:0007611, *p* = .0226).

**Table 3.** Enrichment analysis results for ALSPAC - Read by elastic net model.

| **SNP - Only Model** |  |  |
| --- | --- | --- |
| **Term name** | **Term ID** | **Adjusted p-value** |
| postsynaptic membrane | GO:0045211 | 0.0001 |
| regulation of neuronal synaptic plasticity | GO:0048168 | 0.0007 |
| amyloid-beta binding | GO:0001540 | 0.0410 |
| peptidyl-lysine deacetylation | GO:0034983 | 0.0422 |
| G protein-coupled receptor complex | GO:0097648 | 0.0131 |
| multicellular organismal process | GO:0032501 | 0.0149 |
| learning or memory | GO:0007611 | 0.0226 |
| synaptic membrane | GO:0097060 | 0.0009 |
| plasma membrane region | GO:0098590 | 0.0012 |
|  | GO:0048169 | 0.0032 |
| regulation of long-term neuronal synaptic plasticity |  |  |
| synapse | GO:0045202 | 0.0038 |
| regulation of synaptic plasticity | GO:0048167 | 0.0064 |
| dendritic spine | GO:0043197 | 0.0077 |
| neuron spine | GO:0044309 | 0.0084 |
| Rap1 signaling pathway | KEGG:04015 | 0.0255 |
| multicellular organism development | GO:0007275 | 0.0396 |
| postsynapse | GO:0098794 | 0.0138 |
| Alcoholism | KEGG:05034 | 0.0164 |
| cell junction | GO:0030054 | 0.0168 |
| system process | GO:0003008 | 0.0236 |
| anatomical structure development | GO:0048856 | 0.0285 |
| system development | GO:0048731 | 0.0349 |
| cognition | GO:0050890 | 0.0455 |
| **SNP + Demographics Model** |  |  |
| **Term name** | **Term ID** | **Adjusted p-value** |
| plasma membrane region | GO:0098590 | 0.0025 |
| system process | GO:0003008 | 0.0088 |
| regulation of plasma membrane bounded cell projection organization | GO:0120035 | 0.0175 |
| regulation of dendrite development | GO:0050773 | 0.0103 |
| animal organ development | GO:0048513 | 0.0319 |
| peptidyl-lysine deacetylation | GO:0034983 | 0.0497 |
| postsynaptic membrane | GO:0045211 | 0.0040 |
| synaptic membrane | GO:0097060 | 0.0226 |
| dendrite development | GO:0016358 | 0.0156 |
| regulation of cell projection organization | GO:0031344 | 0.0205 |
| system development | GO:0048731 | 0.0212 |
| anatomical structure development | GO:0048856 | 0.0264 |
| nervous system development | GO:0007399 | 0.0288 |
| dendrite morphogenesis | GO:0048813 | 0.0429 |

In the SNP-plus model, we found 14 enriched pathways, of which six were selected as drivers. These included the plasma membrane region (GO:0098590, *p* = .0025) and the regulation of dendrite development (GO:0050773, *p* = .0103). These pathways reflect neurobiological processes relevant to cognitive and language development, supporting the interpretation that our pipeline successfully identified biologically meaningful predictors of reading ability.

## Discussion

We examined the genetic contributions to reading ability by integrating confirmatory factor analysis, data imputation, genome-wide association studies (GWAS), multivariate elastic net modeling, and pathway enrichment analysis. We hypothesized that combining (1) a multidimensional reading phenotype and (2) elastic net modeling with traditional genetic approaches would yield more informative genetic markers than using conventional methods alone. We also expected that the identified genetic variants would be overrepresented in biological processes related to brain development.

Our results support these hypotheses. By constructing a latent reading ability score and applying complementary analytic techniques, we addressed several limitations of prior genetic studies on reading. Specifically, we identified one genome-wide significant SNP in the ALSPAC cohort, explained an equal or greater proportion of variance in reading ability than traditional approaches, and linked informative markers to biological pathways involved in brain development and cognitive function. Together, these findings highlight the value of applying integrated, alternative statistical pipelines within small but deeply phenotyped cohorts to advance hypothesis generation in the study of complex traits.

### Impact of Latent Trait on Genome-wide Association

Using a latent trait to represent reading ability allowed us to reduce measurement error and increase statistical power. This approach enabled us to identify one genome-wide significant association (rs181384543) after correcting for multiple comparisons. This result demonstrates that refining the phenotype can enhance the detection of genetic signals, even in relatively small samples.

Our GWAS indicated that the C allele of rs181384543, located on chromosome X within the *PTCHD1-AS* gene (Patched Domain-Containing Protein 1, chrXp22.1), was associated with better general reading ability. Previous research has linked disruptions in *PTCHD1* to autism spectrum disorder and intellectual disability^39–41^. Although no prior studies have associated rs181384543 specifically with reading ability or disability, Doust and colleagues reported that several SNPs implicated in autism and intellectual disability also correlated with self-reported dyslexia. These findings support the possibility of shared genetic mechanisms across neurodevelopmental and cognitive traits.

Despite its statistical significance in the GWAS, rs181384543 was not selected by our elastic net models. This may suggest that the SNP contributes little to multivariate interactions or is redundant when modeled alongside other variants. The lack of selection also highlights how univariate significance does not always translate into predictive value in multivariate models.

This result emphasizes the potential importance of sex-linked alleles in reading development. Given the location of rs181384543 on the X chromosome, future studies should investigate how sex-linked genetic variation influences reading outcomes. To confirm the relevance of this SNP, replication in larger and more diverse cohorts will be necessary.

### Variance Explained of Latent Reading Ability

Previous GWAS studies have reported SNP-based heritability estimates ranging from 0.13 for nonword repetition to 0.26 for nonword reading^18^. Our estimate of SNP-based heritability for latent reading ability 0.23 falls within this range and aligns with prior findings. The elastic net model that included only SNPs explained 12 percent of the variance in latent reading ability, which places it at the lower end of the range found in previous research.

The inclusion of child-level demographic, environmental, and behavioral features to the model increased the variance explained to 33 percent. This substantial improvement highlights the critical role of non-genetic factors—such as maternal education, language skills, and cognitive ability—in shaping reading development. In addition to their direct impact, some of these behavioral features may reflect shared genetic architecture with reading ability. For instance, recent genetic studies have identified overlapping associations between reading skills and broader cognitive or educational traits^9,18,42^. It is likely that features such as vocabulary and receptive language were selected in part due to these shared genetic influences.

Despite the improved explanatory power of the combined model, a considerable portion of variance remains unexplained. This underscores the importance of including a broader range of features in future analyses. In this study, we intentionally limited the number of predictors to demonstrate the utility of the pipeline in a focused and interpretable way. However, future applications could incorporate larger sets of genetic variants and a more diverse array of non-genetic features.

Additional non-genetic features could include nonlinguistic measures, such as finger tapping, rhythm judgement, and visual attention, because previous studies suggest a link between motor abilities,^43^ auditory perception,^44^ and visual abilities^45^ with reading and dyslexia. Expanding the scope of predictors would allow for a more comprehensive understanding of the complex factors that shape reading development.

### Links with Theoretically Relevant Biological Functions

Our enrichment analyses identified several overrepresented biological processes involved in brain development, including those related to learning, memory, and dendritic regulation. These findings are consistent with prior genetic research on reading ability, which has linked structural brain differences to reading performance^46,47^, as well as studies demonstrating genetic correlations between reading and other cognitive traits⁹. Since reading is fundamentally a cognitive skill, we expected biological processes related to brain function and development to emerge in the enrichment results.

Importantly, many of the SNPs driving these results were located in genes previously associated with reading ability and reading disorders. One notable example is the postsynaptic membrane pathway (GO:0045211), which was significantly overrepresented in both the SNP-only and SNP-plus models. This suggests that the biological effects of many informative SNPs may operate within postsynaptic regions of neurons, which play critical roles in learning and synaptic plasticity.

Nine SNPs from the SNP-plus model mapped to this cellular component and were located in genes such as *KIAA0319*, *ATP2C2*, *GRIN2B*, *LIN7A*, *TLN1*, *KCNJ10*, and *TNM2*. Several of these genes—*KIAA0319*,^48,49^ *ATP2C2*,^50^ and *GRIN2B*^51^—have well-established links to reading-related traits. Additionally, *ATP2C2,*^52,53^ and *LIN7A*^54,55^ have been implicated in language impairment, further reinforcing the relevance of our findings.

Because these enrichment results reflect known theoretical frameworks and replicate findings from prior studies, they strengthen the validity of our analytic pipeline. Future research can apply this approach to investigate reading and other complex traits, using enrichment analyses to generate biologically meaningful hypotheses that bridge genotype and phenotype.

### Limitations and Future Directions

This study’s primary strength lies in its use of statistical procedures designed to mitigate power loss due to multiple testing corrections in small genetic cohorts. We also demonstrated that a latent reading ability model with acceptable fit can perform comparably to one with near-perfect fit. The consistency of the latent scores across models suggests that researchers can prioritize theoretical coherence and practicality over statistical perfection, especially when modeling complex traits across diverse datasets. This insight has important implications for replicating genetic findings in studies that use different instruments or populations.

Despite these strengths, the study faced several limitations related to sample size, dataset constraints, and analytic methods. Although ALSPAC is one of the largest available cohorts for studying child behavioral genetics, its size remains modest by genomic standards. Small sample sizes are a common limitation in studies of reading ability and reduce the power to detect subtle genetic effects. Furthermore, because the ALSPAC data were collected in the 1990s, we had no control over the selected behavioral measures or the genotyping arrays. These constraints limited the range and specificity of available features and SNP coverage. As such, our findings should be interpreted as hypothesis-generating rather than conclusive.

Methodological limitations also affected our analyses. We ran genome-wide association (GWA) screening chromosome by chromosome due to computational limitations, rather than across the whole genome simultaneously. This approach may have introduced overcorrection during principal component adjustment for genetic similarity. Future studies should replicate our pipeline using updated methods for computing genetic similarity on a genome-wide scale.

In addition, elastic net models do not rely on traditional significance testing to determine the relevance of predictors. In prior studies using similar methods, researchers often input several thousand SNPs^56^. In contrast, we limited our input to fewer than 150 SNPs per analysis to highlight interpretability and methodological transparency. The exclusion of rs181384543—a genome-wide significant SNP in our GWAS—from the elastic net model confirms that feature selection in this context is driven by joint predictive utility rather than univariate significance. Because elastic nets capitalize on intercorrelations among variables, some selected SNPs exhibited multicollinearity, resulting in singularities in subsequent regression models. Still, most features retained in our models have strong support from existing literature and theory, reinforcing the credibility of our pipeline.

Future research should expand the number of input features, including a wider range of genetic markers and non-genetic variables such as motor skills, rhythm perception, and attention measures. This expansion will help refine models of reading ability and further validate the utility of our pipeline in more diverse contexts.

## Conclusions

This study demonstrates how researchers can leverage relatively small but richly phenotyped cohorts to explore the complex genetic and environmental architecture of reading ability. By integrating traditional GWAS with machine learning techniques such as elastic net modeling, we developed a pipeline that successfully identified informative predictors, replicated known genetic associations, and revealed biologically meaningful pathways.

Our findings reinforce the view that reading ability is a multifactorial trait shaped by both genetic and non-genetic influences. We showed that constructing a latent reading phenotype improves power in traditional GWAS, and that elastic net models can detect relevant patterns even without relying on p-values. These models did not generate arbitrary or spurious results but instead produced findings aligned with current theoretical and empirical knowledge.

This analytic approach offers a flexible framework for hypothesis development that can be applied to other complex traits in developmental science. Future research teams can adapt this pipeline to their own data to deepen our understanding of reading development and its broader cognitive and biological underpinnings. By doing so, researchers can continue to refine models of learning that are grounded in both statistical rigor and developmental relevance.

## Data Availability

Please note that the study website contains details of all the data that are available through a fully searchable data dictionary (http://www.bristol.ac.uk/alspac/researchers/our-data/). Data from this study are available through ALSPAC upon approval by the executive board.

## Acknowledgements

We are extremely grateful to all the families who took part in this study, the midwives for their help in recruiting them, and the entire ALSPAC team, which includes interviewers, computer and laboratory technicians, clerical workers, research scientists, volunteers, managers, receptionists and nurses. Thank you to Xiaonan Liu for his help with data analyses on the F32 project. Thank you to everyone who reviewed or provided feedback on a version of this paper.

## Ethics Approval

We obtained ethical approval for this study from the ALSPAC Ethics and Law Committee and the Local Research Ethics Committee(s) (Arizona State University Institutional Review Board). Consent for biological samples has been collected in accordance with Human Tissue Act (2004). Informed consent for the use of data collected via questionnaires and clinics was obtained from participants following the recommendations of the ALSPAC Ethics and Law Committee at the time.

## Availability of Data and Material

Data from this study are available through ALSPAC upon approval by the executive board.

## Code Availability

Code is available in Supplementary Information and at https://www.medrxiv.org/content/10.1101/2021.08.24.21262573v1.

## Authors Contributions

HSL conceptualized the study under the mentorship of JL and VD, analyzed and interpreted the data, drafted the manuscript, and revised it based on feedback from JL, VD, and JG.

## Footnotes

* Genetic similarity is a more accurately descriptive term for the genetic relatedness to describe similarity between participants and reference panels due do common genetic ancestors. Genetic similarity can describe study population as well as control for genetic background.

