## Supplemental Materials for "Pipeline for Identifying Genetic and Demographic Predictors of Latent Reading Ability: Demonstration in ALSPAC Cohort"

#### Supplementary Information Summary

**Supplemental Methods**. Additional details about recruitment, inclusion and exclusion criteria, behavioral assessments, demographic characteristics, and DNA collection procedures

**Supplemental Code**. All code used for data prep, cleaning, and analysis with comments

**Supplemental Table S1**. Demographic information by cohort for children included in genetic analyses

**Supplemental Table S2**. Reading data for ALSPAC cohort

**Supplemental Table S3**. Robust goodness of fit statistics for latent reading ability models in ALSPAC cohort with complete data

**Supplemental Table S4**. List of SNPs included in SNP-only models

**Supplemental Table S5**. Summary of regression tests of elastic net models for ALSPAC - Read

#### Supplemental Methods

We obtained ethical approval for this study from the ALSPAC Ethics and Law Committee and the Local Research Ethics Committee(s) (Arizona State University Institutional Review Board).

For this study, we used data from 8,071 participants who had participated in at least one of the ALSPAC "Focus at" sessions. Our discovery cohort was the ALSPAC. The inclusion criteria for this study were: (1) no diagnosis of autism spectrum disorder, (2) normal hearing status at Focus at 7, (3) nonverbal intelligence greater than 72 standard score on the Weschler Intelligence Scales for Children (Wechsler et al., 1992), (4) completed at least one reading task at Focus at 7 and Focus at 9, and (5) genetic data. Lastly, for twin-pairs one child was randomly selected for analysis to achieve data independence, which resulted in 186 children being removed from the analysis.

#### Measures

We used behavioral and demographic measures, including reading, language, and nonverbal intelligence measures, collected between the ages of 7 and 9. Reading skill was measured during Focus at 7 and Focus at 9 using a combination of word reading, spelling, and connected text tasks. At Focus at 7 years, children completed the single word reading subtest on the Wechsler Objective Reading Dimensions (Rust et al., 1993) and an experimenter derived spelling task (Bryant et al., 2014). Nonword repetition was assessed at Focus at 8 (Gathercole et al., 1994). At Focus at 9, children completed single word reading, nonword reading (Nunes et al., 2003), and spelling tasks similar to the ones presented during Focus at 7 years, but with new words/items. Additionally, at age 9, children completed the Neale Analysis of Reading Ability (NARA; Neale, 1997), which provided scores for reading rate, accuracy, and reading comprehension. The Wechsler Intelligence Scale for Children (WISC; Wechsler et al. 1992) (Focus at 8) yielded an estimate of nonverbal intelligence.  Receptive language was assessed using the Wechsler Objective Language Dimensions Language Comprehension subtest

(WOLD; Rust, 1996). Vocabulary was measured using the Wechsler Intelligence Scales for Children vocabulary subtest (WISC; Wechsler, Golombok, & Rust, 1992).

We selected biological sex, birth weight, maternal education, child ethnicity, bilingual language status, hearing function, and Attention Deficit Hyperactivity Disorder (ADHD) status, as our demographic measures. Biological sex and birth weight in grams were reported at birth. Maternal education was obtained at 32-weeks' gestation and measures the highest degree the mother had obtained by that point: Vocation, certificate of secondary education, O-levels, A-levels, or College degree. Child's ethnicity was reported by mothers at 32 weeks' gestation. Bilingual language status was obtained via parent report at Focus at 8. Hearing functioning was measured via bone conduction at Focus at 7. ADHD) status was determined at age 7 using parent and teacher questionnaires.

**Genotyping**

ALSPAC samples were genotyped using the Illumina HumanHap550 quad chip genotyping platforms by 23andme, subcontracting the Wellcome Trust Sanger Institute, (Cambridge, UK) and the Laboratory Corporation of America (Burlington, NC, USA). The resulting raw genome-wide data were subjected to standard quality control methods. Individuals were excluded on the basis of gender mismatches; minimal or excessive heterozygosity; disproportionate levels of individual missingness (>3%) and insufficient sample replication (identity by descent (IBD) < 0.8). Population stratification was assessed by multidimensional scaling analysis and compared with Hapmap II (release 22) European descent (CEU), Han Chinese, Japanese and Yoruba reference populations; all individuals with non-European ancestry were removed. SNPs with a minor allele frequency of < 1%, a call rate of < 95%, or evidence for violations of Hardy-Weinberg equilibrium (P < 5E-7) were removed. Cryptic relatedness was measured as a proportion of IBD > 0.1. Related subjects that passed all other quality control thresholds were retained during subsequent phasing and imputation. 9,115 participants and 500,527 SNPs passed these quality control filters. Children's genetic data was then combined with mother's genotyped data for imputation (shared SNPs = 477,482). Quality control filters were applied to the combined data (missingness >1%, ID mismatch, HWE). Haplotypes were estimated using ShapeIT (v2.r644). A phased version of the 1000 genomes reference panel (Phase 1, Version 3) was obtained from the Impute2 reference data repository (phased using ShapeIt v2.r644, haplotype release date Dec 2013). Imputation of the target data was performed using Impute V2.2.2 against the reference panel (all polymorphic SNPs excluding singletons), using all 2186 reference haplotypes (including non-Europeans). This gave 8,237 eligible children available genotype data after exclusion of related subjects using cryptic relatedness measures described previously.

**Statistical Analysis**

**Creating reading ability score**. To create the latent reading ability score, we used confirmatory factor analysis (CFA) to load all of the reading variables onto a single factor. We used lavaan (Rosseel, 2012) to fit and assess the reading ability model. Current practice is to use several model fit criteria instead of relying on a single measure. We assessed model fit using a combination of absolute, parsimonious, and comparative indices of model fit (Byrne, 1989). To determine goodness of fit, we evaluated (a) Tucker-Lewis Index (TLI), (b) root mean square error approximation (RMSEA), and (c) standardized root mean square residual (SRMR). We used the TLI to assess comparative or incremental fit. The TLI is a non-normed fit index that is analogous to the r-squared coefficient, with penalties for added parameters. Like the r-squared coefficient, higher values indicate better fit, with the traditional cutoff value for good fit at 0.90. Our index of parsimony was RMSEA. RMSEA ranges from 0 to 1 with values of < 0.08 representing acceptable fit and values < 0.05 representing close fit (Browne & Cudeck, 1992). We report the 90% confidence interval for the RMSEA and the p-value for the closeness of fit test, which tests the null hypothesis that RMSEA is <= 0.05; this test should result in a nonsignificant p-value. Our index of absolute fit was SRMR, which represents the squared difference between observed and predicted correlations and for which values < 0.08 are considered acceptable.

After assessing the fit of the model, we extracted the lambda values associated with manifest paths that exceeded 0.20. The following equation was used to approximate an individual's reading ability score:

$$Reading\ Ability = \lambda_x z_x + \cdots + \lambda_i z_i$$

wherein each lambda was multiplied by the corresponding z-score for a measure and the products were summed together. By using this method, we can approximate the latent construct of reading ability instead of relying on a single reading measure. After approximating the reading ability, we assessed the distribution of scores and normalized if necessary. We used the reading ability score as the phenotype for SNP screening and as the outcome variable for the elastic net model.

**SNP screening**. To constrain the high-dimensionality of the dataset, we selected SNPs using two methods: SNP selection based on review of prior literate and genome-wide association (GWA). We reviewed candidate gene, GWA, and family association studies from 1990 to 2019 for reading disability. We identified 47 SNPs that were reported at least two times to have an association with reading disability; however, we dropped two SNPs (rs93434 and rs454942) due to missingness in the ALSPAC sample.

***GWAS***. GWA was completed in PLINK2 (Chang et al., 2015) and performed chromosome-by-chromosome based on criteria used in prior studies (Lee et al., 2019). We used the standard settings in PLINK. Genetic similarity [*] was controlled for by using the top two principal components. We selected up to 100 SNPs based on uncorrected *p*-values and SNPs

---

[*] Genetic similarity is a more accurately descriptive term for the genetic relatedness to describe similarity between participants and reference panels due do common genetic ancestors. Genetic similarity can describe study population as well as control for genetic background.

with an FDR-BH of 0.1 or less to be included in the subsequent multivariate modeling. See Appendix # for commented code used for all data analysis.

**Machine learning**. There are many options for machine learning. We selected a procedure that would allow for correlated features, more features than subjects, multiple data types, and multiple feature selection. We performed multivariate modeling using an elastic net model to link reading ability with the SNPs that survived the screening step, as well as demographic, environmental, and behavioral covariates. An elastic net is a regularized regression model that enables simultaneous feature selection (in our case, variables are the SNPs) in a high-dimensional setting (Fan & Lv, 2008; Zou, 2006). It adds two regularization terms to the loss function of an ordinary regression model: one L1-norm regularization whose effect is to force the regression coefficients of small effects to be exactly zero, thus enabling feature (e.g., SNP) selection; a second L2-norm regularization term insures highly correlated SNPs are selected. There are two tuning parameters corresponding to the two regularization terms to balance with the loss function. Tuning parameter selection is typically done using cross validation (see below).

We used 5-fold cross-validation to determine the best tuning parameters. In this process, the sample is split into five random groups, four of which are used to train the model and one for testing. This splitting repeats until every “fold” has served as the test set. Cross-validation was performed 10 times to select tuning parameters. After the best tuning parameters were identified, the model was refit using all the data to generate coefficients.

In addition to SNP only modes, we ran SNP plus demographic feature models. For ALSPAC, we included nonverbal IQ, vocabulary, receptive language score, ADHD status, birth weight, bilingual language status, and mother’s highest education in the multivariate model. For GRaD, we selected ADHD status, Hispanic/Latino status, child’s race, mother’s highest education (either birth or, if relevant, adoptive), and vocabulary. However, for GRaD, we were only able to use child’s race and vocabulary score (measured by Peabody Picture Vocabulary), due to large missingness for the other variables (ADHD missing = 266, Hispanic/Latino missing = 138, mother's education missing = 215). We conducted this analysis to determine the impact of SNPs on reading when controlling for demographic, environmental, and behavioral contributions.

**Pathway enrichment and network analysis**. We mapped informative SNPs from the elastic net to genes using g:SNPense on g:Profiler (Reimand et al., 2016). After mapping SNPs to genes, we performed enrichment analysis using g:GOSt on g:Profiler. g:Profiler was selected over similar tools because recent comparisons of the available tools showed that g:Profiler has the most up-to-date repository of pathways and draws from multiple curated sources (e.g., KEGG, Reactome).

**Supplemental Code**

All code used for data prep, cleaning, and analysis with comment.

```
# Generate latent reading variable and add to phenotype file
## Clean ALSPAC data
## Import data
alspac <- read_csv("~/B2569/Lancaster_23August2017.csv")

alspac[alspac < 0] <- NA

## import lists of variable names

read_vars <- read_excel(here("B2569/Dictionary Lancaster.xls"), sheet = "read_vars")

read_vars_names <- read_vars$label
read_vars <- read_vars$variable_name

behav_vars <- read_excel(here("B2569/Dictionary Lancaster.xls"), sheet = "behav_vars")

behav_vars_names <- behav_vars$label
behav_vars <- behav_vars$variable_name

## Create genetic ID

alspac <- alspac %>%
  mutate(gene_id = paste(cidB2569, qlet, sep =""))

## New datasets: alspac_qual & alspac_disqual
### Qualifying children (no hearing loss, no intellectual disability based on full scale, no ASD)

alspac_qual <- alspac %>%
  filter(f7hs035 == 1 | is.na(f7hs035)) %>%
  filter(f8ws112 > 72 | is.na(f8ws112)) %>%
  filter(ku360 != 2 | is.na(ku360))

## New dataset: alspac_qual (Remove 1 twin from the database)

alspac_qual <- alspac_qual %>%
  filter(qlet == "A")

## Data transformations in alspac_qual
alspac_qual <- alspac_qual %>%
  mutate_each(funs(z = scale(.)), one_of(read_vars))

## New dataset: alspac_inall (In all focus sessions)

alspac_inall <- alspac_qual %>%
  mutate(in_all = if_else(in_f07 == 1 & in_f08 == 1 & in_f09 ==1, 1, 0)) %>%
  filter(in_all == 1)

## New datasets: alspac_comp (Complete data only_
```

```
alspac_comp <- alspac_inall %>%
  filter(complete.cases(alspac_inall[ ,behav_vars]))

## Latent reading ability model
reading <- 'reading =~ f7ws076a_z + f7ws117_z + f9mw032_z + f9mw062_z + f9mw098_z +
f9sn705_z + f9sn706_z + f9sn707_z'

fit_read <- cfa(reading, data = alspac_comp, estimator = "MLR", std.lv = TRUE)

summary(fit_read, fit.measures = TRUE, rsquare = TRUE, standardized = TRUE)

parameterEstimates(fit_read, standardized=TRUE) %>%
  filter(op == "=~") %>%
  select('Latent Factor'=lhs, Indicator=rhs, B=est, SE=se, Z=z, 'p-value'=pvalue, Beta=std.all)
%>%
  kable(digits = 3, format="pandoc", caption="Factor Loadings Measurement Model")

reliability(fit_read)

modindices(fit_read)

## extract factor loadings > .20

ro_lambda <- inspect(fit_read, what = "std")$lambda

## mutliple z-scores by factor loading, sum, and transform to z-score

alspac_comp <- alspac_comp %>%
  select(cidB2569, f7ws076a_z, f7ws117_z, f9mw032_z, f9mw062_z, f9mw098_z, f9sn705_z,
f9sn706_z, f9sn707_z) %>%
  mutate(fac_read_loads = ro_lambda[1] * f7ws076a_z +
           ro_lambda[2] * f7ws117_z +
           ro_lambda[3] * f9mw032_z +
           ro_lambda[4] * f9mw062_z +
           ro_lambda[5] * f9mw098_z +
           ro_lambda[6] * f9sn705_z +
           ro_lambda[7] * f9sn706_z +
           ro_lambda[8] * f9sn707_z) %>%
  mutate(fac_read_comp = scale(fac_read_loads))

## CFA with correlated error terms
reading_mod8 <- 'reading =~ f7ws076a_z + f7ws117_z + f9mw032_z + f9mw062_z +
f9mw098_z + f9sn705_z + f9sn706_z + f9sn707_z
f7ws076a_z ~~ f7ws117_z
f9sn706_z ~~ f9sn707_z
f9mw032_z ~~ f9mw062_z
f7ws117_z ~~ f9mw098_z
f9mw032_z ~~ f9mw098_z
f9mw062_z ~~ f9mw098_z
f9sn705_z ~~ f9sn707_z
```

```
f9mw032_z ~~ f9sn707_z'

fit_read_mod8 <- cfa(reading_mod8, data = alspac_comp, estimator = "MLR", std.lv = TRUE)

summary(fit_read_mod8, fit.measures = TRUE, rsquare = TRUE, standardized = TRUE)

parameterEstimates(fit_read_mod8, standardized=TRUE) %>%
  filter(op == "=~") %>%
  select('Latent Factor'=lhs, Indicator=rhs, B=est, SE=se, Z=z, 'p-value'=pvalue, Beta=std.all)
%>%
  kable(digits = 3, format="pandoc", caption="Factor Loadings Measurement Model")

## compare base score to model with correlated errors

## extract factor loadings > .20

ro_lambda <- inspect(fit_read_mod8, what = "std")$lambda

## extract the scores & link to participant id

case_ids <- inspect(fit_read_mod8, "case.idx")
pred1 <- predict(fit_read_mod8)
pred1 <- data.frame(pred1, id = case_ids)
alspac_comp <- cbind(alspac_comp, pred1["reading"])

## compare base score to mod 8 score

t.test(alspac_comp$fac_read_comp, alspac_comp$reading, paired = TRUE, alternative =
"two.sided")

# QC ALSPAC raw genetic files
cd chr/
mkdir work_dir/problematic_variants

pwd

for i in {01..22}
do

#use plink to convert to pgen format
plink2 --bgen data_chr${i}.bgen ref-first --sample dataFac.sample --missing-code -9 --keep
unrelated_children.txt --memory 3000 --make-pgen --out work_dir/"ALSPAC_raw_chr${i}"

#set var ids using chr pos ref alt format
plink2 --pfile work_dir/"ALSPAC_raw_chr${i}" --set-all-var-ids @:#\$r\$a --new-id-max-allele-len
23 truncate --make-pgen --out work_dir/"ALSPAC_raw_chr${i}"

#use plink to find problematic variants, including duplicates and split multiallelic var
plink2 --pfile work_dir/"ALSPAC_raw_chr${i}" --rm-dup list --out
work_dir/problematic_variants/"ALSPAC_raw_chr${i}_noext"
```

```
#check if problematic variants were found
if [[ -s work_dir/problematic_variants/"ALSPAC_raw_chr${i}_noext.rmdup.mismatch" ]]; then

#exclude problematic variants from the original pgen file
plink2 --pfile work_dir/"ALSPAC_raw_chr${i}" --exclude
work_dir/problematic_variants/"ALSPAC_raw_chr${i}_noext.rmdup.mismatch" --make-pgen --
out work_dir/"ALSPAC_chr${i}_clean"
else

#no problematic variants found, copy original file
cp work_dir/"ALSPAC_raw_chr${i}.pgen" work_dir/"ALSPAC_chr${i}_clean.pgen"
cp work_dir/"ALSPAC_raw_chr${i}.psam" work_dir/"ALSPAC_chr${i}_clean.psam"
cp work_dir/"ALSPAC_raw_chr${i}.pvar" work_dir/"ALSPAC_chr${i}_clean.pvar"
fi

done

# Generate PCs chr by chr

cd chr/

for i in {01..21}
do
  echo "Processing Chromosome ${i}"

# Calculating PCs:
# Keep only SNPs with MAF > 10% and prune SNPs, then calculate PCs
plink2 --pfile work_dir/"ALSPAC_chr${i}_clean" --keep unrelated_children.txt --memory 9000 --
mach-r2-filter .95 1 --maf 0.1 --indep-pairwise 1500 150 0.2 --pca --out
work_dir/"ALSPAC_chr${i}_clean_PCA"

done

# Run GWAS with top 3 PCs chr by chr

cd chr/

for i in {01..22}
do
  echo "Processing Chromosome ${i}"

# step 1: run glm with cleaning
plink2 --pfile work_dir/"ALSPAC_chr${i}_clean" --keep unrelated_children.txt --maf 0.1 --indep-
pairwise 1500 150 .2 --mach-r2-filter .95 1 --covar
work_dir/"ALSPAC_chr${i}_clean_PCA.eigenvec" --covar-name PC1, PC2, PC3 --glm hide-
covar --out work_dir/"ALSPAC_chr${i}"

# step 2: down stream

#number of lines with NA present
```

```
grep -E "NA" -c work_dir/"ALSPAC_chr${i}.fac_read_comp.glm.linear" >>
work_dir/"ALSPAC_chr${i}_fac_qc"

# step 3: clean copy of results

grep -v "NA" work_dir/"ALSPAC_chr${i}.fac_read_comp.glm.linear" >
work_dir/"ALSPAC_chr${i}.fac_read_compC.glm.linear"

# step 4: prep QQ-plot file

#awk '{if (NR>1) print $1,$3,$15}' work_dir/"ALSPAC_chr${i}.cont.assoc.linear" >
work_dir/"ALSPAC_chr${i}.plot.adclean.cont.linear.txt"

# step 5: top 100 before adjustment

sort -k15 -n work_dir/"ALSPAC_chr${i}.fac_read_compC.glm.linear" >
work_dir/"ALSPAC_chr${i}_fac_read_compC_sort"
head -n 101 work_dir/"ALSPAC_chr${i}_fac_read_compC_sort" >
work_dir/"ALSPAC_chr${i}_top100"

# step 6: p-value adjustment

plink2 --adjust-file work_dir/"ALSPAC_chr${i}.fac_read_comp.glm.linear" test=ADD --out
work_dir/"ALSPAC_chr${i}_fac"

# step 7: significant results after adjustment

#sort by pvalue for FDRBH
sort -k15 -n work_dir/"ALSPAC_chr${i}_fac.adjusted" >
work_dir/"ALSPAC_chr${i}_fac_adj_sorted.txt"

#extract lines that are nominal significant based on FDRBH
awk ' $15 <= 0.05 ' work_dir/"ALSPAC_chr${i}_fac.adjusted" >
work_dir/"ALSPAC_chr${i}_fac_sig_results.txt"

# step 7: make chr directory

mkdir work_dir/ALSPAC_chr${i}_fac/

# step 8: mv all files to new directory

mv work_dir/"ALSPAC_chr${i}*" ALSPAC_chr${i}_fac/

done

# SNP-based heritability prep and analysis code

## use ldsc 1kg-p3 grCh37 bim file

wget "https://zenodo.org/record/7768714/files/1000G_Phase3_plinkfiles.tgz?download=1" \
-O 1000G_Phase3_plinkfiles.tgz
```

```
tar -xvzf 1000G_Phase3_plinkfiles.tgz

# concatenate all 22 chunks into a single bim file

cat 1000G.EUR.QC.*.bim > 1000G.EUR.QC.all.bim

module load r-4.4.2-gcc-12.1.0
library(data.table)
bim <- fread("1000G.EUR.QC.all.bim",
            col.names = c("chr","rsid","cm","bp","a1_ref","a2_ref"))

### merge gwas with ref bim file to add rsid's

library(data.table)
library(dplyr)

# File paths
gwas_file <- "Input_ldsc_ALSPC.txt"
bim_file <- "1000G.EUR.QC.all.bim"
output_file <- "Input_ldsc_ALSPC_with_rsid.txt"

# Load GWAS data
gwas <- fread(gwas_file)

# Load BIM file
bim <- fread(bim_file, col.names = c("chr", "rsid", "cm", "bp", "a1_ref", "a2_ref"))

# Merge logic: match on chr and bp and alleles (both orientations)
# Merge with direct match
merged1 <- gwas %>%
  inner_join(bim, by = c("chr", "bp", "a1" = "a1_ref", "a2" = "a2_ref"))

# Merge with flipped alleles
merged2 <- gwas %>%
  inner_join(bim, by = c("chr", "bp", "a1" = "a2_ref", "a2" = "a1_ref"))

# Combine both
merged <- bind_rows(merged1, merged2) %>% distinct()

# Drop duplicates, select final columns
final <- merged %>%
  select(rsid, chr, bp, a1, a2, n, beta, se, pval)

# Save to output
fwrite(final, output_file, sep = "\t")

## munge the summary stats file first
./munge_sumstats.py \
--sumstats Input_ldsc_ALSPC_with_rsid.txt \
--N 8071 \
```

```
--out ALSPAC \
--chunksize 500000 \
--merge-alleles w_hm3.snplist

### compute heritability estimates (h2)

## heritability estimates

./ldsc.py \
--h2 ALSPAC.sumstats.gz \
--ref-ld-chr eur_w_ld_chr/ \
--w-ld-chr eur_w_ld_chr/ \
--out alspac_h2

# Add rs-IDs using biomaRt for downstream processing for elastic net modeling

## R code to add rs-ids to combined summary stats file

if (!require("BiocManager", quietly = TRUE))
   install.packages("BiocManager")
BiocManager::install(version = "3.20")

BiocManager::install(c("GenomicFeatures", "AnnotationDbi"))

BiocManager::install("biomaRt")

library(biomaRt)

mart <- useMart("ENSEMBL_MART_SNP", "hsapiens_snp",
"https://feb2014.archive.ensembl.org")

 all_top100 <- all_top100 %>%
         mutate(coords = paste(CHROM, POS, POS, sep = ":")

coords_df <- all_top100$coords

## Submit query

rsids_top100 <- getBM(attributes = c('refsnp_id', 'chr_name', 'chrom_start', 'allele'),
     filters = c('chromosomal_region'),
     values = coords_df,
     mart = mart)

## Join and write out

all_top100 <- all_top100 %>%
         left_join(rsids_top100, by = c("CHROM" = "chr_name", "POS" = "chrom_start"))

write.table(all_top100, file = "all_top100_rsids.txt", sep = "\t", row.names = FALSE, quote =
FALSE)
```

#### getting chr position for prev lit snps

```
m <- biomaRt::getBM(attributes = c('refsnp_id', 'chr_name', 'chrom_start', 'allele'), #what we are
getting from the query
                    filters = c('snp_filter'), #where to look in the mart
                    values = snp_filter,  #what to look for
                    mart = mart) #the mart to use
colnames(m) <- c("SNP", "CHR", "BP", "Allele")

m <- m[-c(4:8)] #dropped allele column

m2 <- m[-c(4:8),] # dropped duplicated rows
```

#### Join with top 100 SNPs and add coords in correct format

```
fac_input <- top100 %>%
  select(refsnp_id, CHROM, POS) %>%
  rename(SNP = refsnp_id,
         CHR = CHROM,
         BP = POS) %>%
  rbind(m2)

fac_input <- fac_input %>%
          mutate(coords = paste0("chr",CHR, ":", BP, "-", BP))
```

#### write out

```
write.table(fac_input, file = "fac_input.txt", sep = "\t", row.names = FALSE, quote = FALSE)
```

### Create genotype file for elastic net modeling

#### script to pull out snp genotype probabilities from all chr files for latent reading ability
#### the first line of code goes through all the chr files and extracts relevant SNPs from the chr bgen file
#### the second line of code creates the necessary bgi file for further processing
#### both files for each chr must be in the extract file BEFORE concatenation can be done!!!

```
cd qctool/
echo $1
module load gcc/8.2.0

./qctool_v2.2.0 -g ~/Desktop/F32_ALSPAC/data/chr/data_chr$1.bgen -s
/~/Desktop/F32_ALSPAC/data/chr/data.sample -incl-rsids
/~/Desktop/F32_ALSPAC/data/chr/fac_rsids_only.txt -og
~/Desktop/F32_ALSPAC/data/chr/work_dir/fac_extracts/chr$1_snp_ra.bgen

bgenix -g ~/Desktop/F32_ALSPAC/data/chr/work_dir/fac_extracts/chr$1_snp_ra.bgen -index
```

### Elastic net modeling code

#### Import genotypes

```
fac_snp_g <- read_tsv(here("fac_snp_g_20200311.txt"))

setnames(fac_snp_g, "X1", "gene_id", skip_absent = T)

## Import list of unrelated

unrelated_children <- read_tsv(here("data/chr/unrelated_children.txt"), col_names = FALSE,
trim_ws = TRUE)

## Data transformations

alspac_comp <- alspac_comp %>%
  mutate_each(funs(z = scale(.)), c(f8at062, f8sl040, f8ws053, kz030, bestgest, f8ws111))

## set up small data frames for elastic net modeling

alspac_fac_read <- alspac_inall %>%
  select(gene_id, fac_read_comp)

fac_lasso <- fac_snp_g %>%
  left_join(alspac_fac_read, by = "gene_id") %>% # join genotype data with demographic and
behavioral data
  filter(gene_id %in% unrelated_children$X1) %>% # include only unrelated children
  filter(!is.na(fac_read_comp) & fac_read_comp != -9) # remove missing data

## run lasso models - SNP only model first
set.seed(1234)

fac_lasso_full=fac_lasso[!is.na(fac_lasso$fac_read_comp),]
X_fac=as.matrix(fac_lasso_full[,-c(1,150)])
y_fac=as.matrix(fac_lasso_full[,150])
alpha=1:10/10
lassolist=vector("list", 10)
cv=rep(0,10)
for(i in 1:10){
  rdlasso=cv.glmnet(X_fac, y_fac, alpha=alpha[i], nfold=5)
  cv[i]=min(rdlasso$cvm)
  lassolist[[i]]=glmnet(X_fac, y_fac, alpha=alpha[i],lambda=rdlasso$lambda.min)
}
index=which.min(cv)

e <- as.matrix(lassolist[[index]]$beta)
e <- as.data.frame(e)
e$s0 <- abs(e$s0)

e<-e %>% rownames_to_column('new_column')
e<-filter_at(e, vars(-new_column), any_vars(. > 0.019))
e<-e %>% column_to_rownames('new_column')

preds_e <- row.names(e)
fac_lasso_selected <- fac_lasso_full %>%
```

```
  select(fac_read_comp, one_of(preds_e))

fac_lin_reg <- lm(fac_read_comp ~ ., data = fac_lasso_selected)
fac_lin_null <- lm(fac_read_comp ~ 1, data = fac_lasso_selected)
summary(fac_lin_reg)

anova(fac_lin_null, fac_lin_reg)

## SNP+demographics model
alspac_fac_snp_deg=alspac_inall[,c("gene_id", "fac_read_comp", "f8ws111", "kz030", "c645a", "f8ws053", "f8sl040", "f8sl201", "kz021.x", "c804")]


fac_lasso_snp_deg <- fac_snp_g %>%
  left_join(alspac_fac_snp_deg, by = "gene_id") %>%
  filter(gene_id %in% unrelated_children$X1) %>%
  filter(!is.na(fac_read_comp) & fac_read_comp != -9)

a=fac_lasso_snp_deg$f8ws111
fac_lasso_snp_deg$f8ws111=(a-mean(a, na.rm=TRUE))/sd(a, na.rm=TRUE)
a=fac_lasso_snp_deg$kz030
fac_lasso_snp_deg$kz030=(a-mean(a, na.rm=TRUE))/sd(a, na.rm=TRUE)
a=fac_lasso_snp_deg$c645a
fac_lasso_snp_deg$c645a=(a-mean(a, na.rm=TRUE))/sd(a, na.rm=TRUE)
a=fac_lasso_snp_deg$f8ws053
fac_lasso_snp_deg$f8ws053=(a-mean(a, na.rm=TRUE))/sd(a, na.rm=TRUE)
a=fac_lasso_snp_deg$f8sl040
fac_lasso_snp_deg$f8sl040=(a-mean(a, na.rm=TRUE))/sd(a, na.rm=TRUE)

fac_lasso_full_deg=na.omit(fac_lasso_snp_deg)                    ####eliminate NA
X_deg=as.matrix(fac_lasso_full_deg[,-c(1,150)])                 ####predictor
y_deg=as.matrix(fac_lasso_full_deg[,150])                       ####response
alpha=0:10/10
lassolist=vector("list", 11)
cv=rep(0,11)
for(i in 1:11){
  faclasso=cv.glmnet(X_deg, y_deg,  alpha=alpha[i], nfold=5)
  cv[i]=min(rdlasso$cvm)
  lassolist[[i]]=glmnet(X_deg, y_deg, alpha=alpha[i],lambda=rdlasso$lambda.min)
}
index=which.min(cv)


faclasso=cv.glmnet(X_fac, y_fac, nfold=10)
faclasso1=glmnet(X_fac, y_fac, lambda=faclasso$lambda.min)
faclasso2=glmnet(X_fac, y_fac, lambda=faclasso$lambda.1se)

f <- as.matrix(lassolist[[index]]$beta)
f <- as.data.frame(f)
f$s0 <- abs(f$s0)
```

```
f<-f %>% rownames_to_column('new_column')
f<-filter_at(f, vars(-new_column), any_vars(. > 0.019))
f<-f %>% column_to_rownames('new_column')

preds_f <- row.names(f)
fac_lasso_dem_selected <- fac_lasso_full_deg %>%
  select(fac_read_comp, one_of(preds_f))

fac_lasso_dem_preds_b <- fac_lasso_full_deg %>%
  select(fac_read_comp, one_of(preds_e))

fac_dem_lin_reg <- lm(fac_read_comp ~ ., data = fac_lasso_dem_selected)
fac_dem_lin_null <- lm(fac_read_comp ~ 1, data = fac_lasso_dem_selected)
fac_dem_lin_b <- lm(fac_read_comp ~ ., data = fac_lasso_dem_preds_b)
summary(fac_dem_lin_reg)
summary(fac_dem_lin_b)
anova(fac_dem_lin_null, fac_dem_lin_reg)
anova(fac_dem_lin_b, fac_dem_lin_reg)

### Correlations reading and demographic features
cormat <- fac_lasso_full_deg %>%
  dplyr::select(fac_read_comp, kz021.x, c804, f8sl040, f8ws111, f8ws053, c645a, kz030,
f8sl201) %>%
  do(as.data.frame(round(cor(., method = "pearson", use = "pairwise.complete.obs"), 2)))

# Get lower triangle of the correlation matrix
  get_lower_tri<-function(cormat){
    cormat[upper.tri(cormat)] <- NA
    return(cormat)
  }
  # Get upper triangle of the correlation matrix
  get_upper_tri <- function(cormat){
    cormat[lower.tri(cormat)]<- NA
    return(cormat)
  }

upper_tri <- get_upper_tri(cormat)
melted_cormat <- melt(as.matrix(upper_tri), na.rm = TRUE)

## Figure 2 Plot code

## Reading and biological sex

sex_plot <- ggplot(fac_lasso_full_deg, aes(y = fac_read_comp, x = as.factor(kz021.x))) +
  geom_boxplot() +
  stat_summary(fun=mean, geom="point", shape=23, size=4, position=position_dodge(.75)) +
  scale_x_discrete(name = "Biological Sex", labels=c("1" = "Male", "2" = "Female")) +
  labs(y = "Latent Reading Ability") +
  theme_classic()

sex_comp <- fac_lasso_full_deg %>%
```

```
  group_by(kz021.x) %>%
  summarise(mean_read = mean(fac_read_comp, na.rm = T),
            sd_read = sd(fac_read_comp, na.rm = T))

sex_comp

t.test(log(fac_read_comp) ~ kz021.x, data = fac_lasso_full_deg)

### Reading and Parent reported Ethnicity/Race

race_plot <- ggplot(fac_lasso_full_deg, aes(y = fac_read_comp, x = as.factor(c804))) +
  geom_boxplot() +
  stat_summary(fun=mean, geom="point", shape=23, size=4, position=position_dodge(.75)) +
  scale_x_discrete(name = "Ethnicity/Race", labels=c("1" = "White", "2" = "Non-white")) +
  labs(y = "Latent Reading Ability") +
  theme_classic()

eth_comp <- fac_lasso_full_deg %>%
  group_by(c804) %>%
  summarise(mean_read = mean(fac_read_comp, na.rm = T),
            sd_read = sd(fac_read_comp, na.rm = T))

eth_comp

t.test(log(fac_read_comp) ~ c804, data = fac_lasso_full_deg)

### Reading and receptive language

receplang_plot <- ggplot(fac_lasso_full_deg, aes(y = fac_read_comp, x = f8sl040)) +
  geom_point() +
  stat_smooth() +
  labs(y = "Latent Reading Ability", x = "WOLD Receptive Language \n (z-score)") +
  theme_classic()

### Reading and bilingual status

biling_plot <- ggplot(fac_lasso_full_deg, aes(y = fac_read_comp, x = as.factor(f8sl201))) +
  geom_boxplot() +
  stat_summary(fun=mean, geom="point", shape=23, size=4, position=position_dodge(.75)) +
  scale_x_discrete(name = "Child Uses Other Language", labels=c("1" = "Yes", "2" = "No", "9" =
"Unknown")) +
  labs(y = "Latent Reading Ability") +
  theme_classic()

### Reading and scaled preferred birthweight

birthw_plot <- ggplot(fac_lasso_full_deg, aes(y = fac_read_comp, x = kz030)) +
  geom_point() +
  stat_smooth() +
  labs(y = "Latent Reading Ability", x = "Scaled Birthweight \n (z-score)") +
  theme_classic()
```

```
### Reading and NVIQ

nviq_plot <- ggplot(fac_lasso_full_deg, aes(y = fac_read_comp, x = f8ws111)) +
  geom_point() +
  stat_smooth() +
  labs(y = "Latent Reading Ability", x = "WISC Performance IQ \n (z-score)") +
  theme_classic()

### Reading and WISC Vocab

vocab_plot <- ggplot(fac_lasso_full_deg, aes(y = fac_read_comp, x = f8ws053)) +
  geom_point() +
  stat_smooth() +
  labs(y = "Latent Reading Ability", x = "WISC Vocabulary \n (z-score)") +
  theme_classic()


### Reading and highest level of maternal education

momed_plot <- ggplot(fac_lasso_full_deg, aes(y = fac_read_comp, x = as.factor(c645a.y))) +
  geom_boxplot() +
  stat_summary(fun=mean, geom="point", shape=23, size=4, position=position_dodge(.75)) +
  scale_x_discrete(name = "Mother's Highest Education",
                labels=c("0" = "None",
                         "1" = "CSE",
                         "2" = "Vocational",
                         "3" = "O Level",
                         "4" = "A Level",
                         "5" = "Degree")) +
  labs(y = "Latent Reading Ability") +
  theme_classic()

### Combine plots

geneb_plots <- ggarrange(sex_plot, race_plot, biling_plot, momed_plot,
                         vocab_plot, nviq_plot, receplang_plot, birthw_plot,
                         nrow = 4, ncol = 2,
                         labels = "AUTO",
                         font.label = list(size = 10),
                         align = "hv",
                         common.legend = TRUE,  # Enable common legend
                          legend = "bottom")

geneb_plots_ann <- annotate_figure(geneb_plots,
               top = text_grob("Relationships between Latent Reading Ability and Demographic and
Behavioral Variables in ALSPAC cohort (n = 3,254)", color = "black",
                               size = 12))
geneb_plots_ann
```

Supplemental Table S1

*Demographic information by cohort for children included in genetic analyses*

| | |
|---|---|
| **N** | 7977 |
| **Age** [*a] | --- |
| **Male** [*b] | 4036 |
| **Race/Ethnicity** | |
| White | 4965 |
| Non-white | 185 |
| African American | --- |
| Hispanic | --- |
| Missing | 445 |
| **ADHD** | 78 |
| **Bilingual** | 136 |
| **Birthweight (grams)** | n = 4014 m = 3444.63 (521.95) |
| **Mother's education** | |
| CSE | 568 |
| Vocational | 427 |
| O Levels | 1819 |
| A Levels | 1479 |
| Degree | 932 |
| Less than 7 years | --- |
| 7 - 9 years | --- |
| 10 -11 years | --- |
| High school diploma/GED | --- |
| Associate's/Trade/Business | --- |
| Bachelor's | --- |
| Professional or Advanced degree | --- |
| Missing | 370 |
| **Vocabulary** | |
| WISC - Vocabulary | n = 4185, m = 11.52 (4.29) |
| PPVT | --- |
| **Receptive language** | n = 4205 m = 7.60 (1.91) |

Supplemental Table S2

*Reading data for ALSPAC cohort*

| | |
|---|---|
| **Phonological Awareness** | |
| Phoneme deletion | n = 5513, m = 20.78 (9.22) |
| Nonword repetition | n = 5537, m = 7.36 (2.45) |
| **Single Word Reading** | |
| Single word reading at 7 | n = 5518 m = 29.12 (8.87) |
| Single word reading at 9 | n = 5546 m = 7.73 (2.23) |
| Nonword reading at 9 | n = 5542 m = 5.36 (2.43) |
| **Spelling** | |
| Spelling at 7 | n = 5445 m = 26.79 (12.23) |
| Spelling at 9 | n = 5537 m = 10.48 (3.27) |
| **Connected Text** | |
| NARA - Rate | n = 4972 m = 106.15 (12.25) |
| NARA - Accuracy | n = 4893 m = 105.06 (13.19) |
| NARA - Comprehension | n = 4983 m = 101.32 (11.41) |

n = Participants with data, m = Mean (standard deviation)

Supplemental Table S3

*Robust goodness of fit statistics for latent reading ability models in ALSPAC cohort with complete data*

| | ALSPAC | | | |
|---|---|---|---|---|
| **Model** | **Chi-square** | **TLI** | **RMSEA** | **SRMR** |
| Base | 1307.97, df = 27, p < .001 | 0.925 | 0.124, 95% CI [0.118, 0.131] *p* = < .001 | 0.033 |
| Best fitting (8 correlated errors) | 174.73, df = 12, p < .001 | 0.982 | 0.066, 95% CI [0.058, .075], p = .002 | 0.015 |

Supplemental Table S4

*List of SNPs included in SNP-only models*

| SNP | Source | Chromosome | Position |
|---|---|---|---|
| rs12118079 | ALSPAC - Read | 1 | 118238483 |
| rs1984411 | ALSPAC - Read | 1 | 9270072 |
| rs12404907 | ALSPAC - Read | 1 | 9270511 |
| rs12086013 | ALSPAC - Read | 1 | 159983651 |
| rs2904122 | ALSPAC - Read | 1 | 23059784 |
| rs7577992 | ALSPAC - Read | 2 | 41725706 |
| rs4563279 | ALSPAC - Read | 2 | 41752612 |
| rs7586603 | ALSPAC - Read | 2 | 41752842 |
| rs6544443 | ALSPAC - Read | 2 | 41751190 |
| rs4952486 | ALSPAC - Read | 2 | 41748171 |
| rs7586252 | ALSPAC - Read | 2 | 7425507 |
| rs13408091 | ALSPAC - Read | 2 | 170657914 |
| rs11901326 | ALSPAC - Read | 2 | 238261509 |
| rs4245775 | ALSPAC - Read | 2 | 41755591 |
| rs6758123 | ALSPAC - Read | 2 | 170664512 |
| rs617829 | ALSPAC - Read | 2 | 240142864 |
| rs7624403 | ALSPAC - Read | 3 | 63970077 |
| rs2630806 | ALSPAC - Read | 3 | 21892251 |
| rs200564558 | ALSPAC - Read | 3 | 21892251 |
| rs1486369 | ALSPAC - Read | 3 | 21887786 |
| rs2358932 | ALSPAC - Read | 3 | 22756005 |
| rs832197 | ALSPAC - Read | 3 | 63884292 |
| rs59191864 | ALSPAC - Read | 4 | 142241253 |
| rs55703414 | ALSPAC - Read | 4 | 160075956 |
| rs7681750 | ALSPAC - Read | 4 | 160084403 |
| rs41508146 | ALSPAC - Read | 4 | 142244179 |
| rs62327609 | ALSPAC - Read | 4 | 142240955 |
| rs7436963 | ALSPAC - Read | 4 | 142265477 |
| rs1369973 | ALSPAC - Read | 4 | 142233546 |
| rs6537043 | ALSPAC - Read | 4 | 142233426 |
| rs36018887 | ALSPAC - Read | 4 | 160087486 |
| rs6536395 | ALSPAC - Read | 4 | 160080870 |
| rs7673630 | ALSPAC - Read | 4 | 142241448 |
| rs13123795 | ALSPAC - Read | 4 | 160085686 |
| rs7685977 | ALSPAC - Read | 4 | 142233434 |
| rs709390 | ALSPAC - Read | 5 | 98329475 |
| rs1508818 | ALSPAC - Read | 5 | 41888912 |
| rs11750607 | ALSPAC - Read | 5 | 41889050 |
| rs1035418 | ALSPAC - Read | 5 | 166971830 |
| rs154205 | ALSPAC - Read | 5 | 97985302 |
| rs416750 | ALSPAC - Read | 5 | 98334750 |
| rs10040066 | ALSPAC - Read | 5 | 98337037 |

| | | | |
|---|---|---|---|
| rs4565239 | ALSPAC - Read | 5 | 98337848 |
| rs35244816 | ALSPAC - Read | 5 | 41901324 |
| rs77524385 | ALSPAC - Read | 5 | 98328974 |
| rs447044 | ALSPAC - Read | 5 | 98331276 |
| rs1472622 | ALSPAC - Read | 5 | 98336453 |
| rs113868271 | ALSPAC - Read | 5 | 98328445 |
| rs112393710 | ALSPAC - Read | 5 | 98328465 |
| rs111871078 | ALSPAC - Read | 5 | 98328470 |
| rs1427079 | ALSPAC - Read | 6 | 104486858 |
| rs67554174 | ALSPAC - Read | 7 | 9717459 |
| rs7798197 | ALSPAC - Read | 7 | 19037661 |
| rs11525821 | ALSPAC - Read | 7 | 9714758 |
| rs67906523 | ALSPAC - Read | 7 | 9717905 |
| rs68056031 | ALSPAC - Read | 7 | 9716998 |
| rs57301765 | ALSPAC - Read | 7 | 19052733 |
| rs10231255 | ALSPAC - Read | 7 | 45174089 |
| rs11767365 | ALSPAC - Read | 7 | 45225854 |
| rs13307587 | ALSPAC - Read | 7 | 13543729 |
| rs4292642 | ALSPAC - Read | 8 | 82873063 |
| rs5025174 | ALSPAC - Read | 8 | 4287495 |
| rs7837231 | ALSPAC - Read | 8 | 82859936 |
| rs10814272 | ALSPAC - Read | 9 | 35715911 |
| rs2295794 | ALSPAC - Read | 9 | 35700150 |
| rs10795024 | ALSPAC - Read | 10 | 3359982 |
| rs7130517 | ALSPAC - Read | 11 | 129760395 |
| rs67311620 | ALSPAC - Read | 11 | 129724031 |
| rs2141786 | ALSPAC - Read | 11 | 129722782 |
| rs56112470 | ALSPAC - Read | 11 | 129753945 |
| rs2303662 | ALSPAC - Read | 11 | 129754668 |
| rs55900082 | ALSPAC - Read | 11 | 129723248 |
| rs67309388 | ALSPAC - Read | 12 | 49685089 |
| rs73144951 | ALSPAC - Read | 12 | 81316742 |
| rs1054442 | ALSPAC - Read | 12 | 49389320 |
| rs7154388 | ALSPAC - Read | 14 | 96169962 |
| rs7158782 | ALSPAC - Read | 14 | 96169131 |
| rs61997099 | ALSPAC - Read | 14 | 104766616 |
| rs231684 | ALSPAC - Read | 17 | 3207447 |
| rs231688 | ALSPAC - Read | 17 | 3207019 |
| rs6502712 | ALSPAC - Read | 17 | 3204033 |
| rs368810 | ALSPAC - Read | 17 | 3236930 |
| rs370361 | ALSPAC - Read | 17 | 3237419 |
| rs371236 | ALSPAC - Read | 17 | 3241329 |
| rs12150649 | ALSPAC - Read | 17 | 3335290 |
| rs2170686 | ALSPAC - Read | 17 | 3288545 |
| rs1067094 | ALSPAC - Read | 17 | 3285471 |
| rs2676612 | ALSPAC - Read | 17 | 3225404 |

| | | | |
|---|---|---|---|
| rs1084906 | ALSPAC - Read | 17 | 3236291 |
| rs1067091 | ALSPAC - Read | 17 | 3240297 |
| rs2676610 | ALSPAC - Read | 17 | 3222813 |
| rs112733297 | ALSPAC - Read | 17 | 3287288 |
| rs804230 | ALSPAC - Read | 17 | 3218752 |
| rs804231 | ALSPAC - Read | 17 | 3220544 |
| rs7220982 | ALSPAC - Read | 17 | 3376682 |
| rs7207788 | ALSPAC - Read | 17 | 3376885 |
| rs230467 | ALSPAC - Read | 17 | 3258491 |
| rs230472 | ALSPAC - Read | 17 | 3266022 |
| rs1468647 | ALSPAC - Read | 18 | 53767627 |
| rs17689314 | ALSPAC - Read | 21 | 26182836 |
| rs181384543 | ALSPAC - Read | X | 22779030 |
| rs333491 | Previous Literature | 3 | 78808850 |
| rs796503097 | Previous Literature | 3 | 134264558 |
| rs111961595 | Previous Literature | 6 | 41147625 |
| rs2274305 | Previous Literature | 6 | 24291203 |
| rs3765502 | Previous Literature | 6 | 24354045 |
| rs793862 | Previous Literature | 6 | 24207200 |
| rs807701 | Previous Literature | 6 | 24273791 |
| rs807724 | Previous Literature | 6 | 24278869 |
| rs7765678 | Previous Literature | 6 | 24330544 |
| rs16889556 | Previous Literature | 6 | 24641605 |
| rs16889506 | Previous Literature | 6 | 24595853 |
| rs699463 | Previous Literature | 6 | 24544903 |
| rs2143340 | Previous Literature | 6 | 24659071 |
| rs4504469 | Previous Literature | 6 | 24588884 |
| rs2038137 | Previous Literature | 6 | 24645943 |
| rs6935076 | Previous Literature | 6 | 24644322 |
| rs2710102 | Previous Literature | 7 | 147574390 |
| rs7782412 | Previous Literature | 7 | 114290415 |
| rs936146 | Previous Literature | 7 | 114294405 |
| rs923875 | Previous Literature | 7 | 113735036 |
| rs1163203 | Previous Literature | 10 | 70554635 |
| rs1079727 | Previous Literature | 11 | 113289182 |
| rs5796555 | Previous Literature | 12 | 13855534 |
| rs1012586 | Previous Literature | 12 | 13855632 |
| rs2268119 | Previous Literature | 12 | 13872634 |
| rs2216128 | Previous Literature | 12 | 13883014 |
| rs2192973 | Previous Literature | 12 | 13896555 |
| rs2289105 | Previous Literature | 15 | 51507508 |
| rs1065778 | Previous Literature | 15 | 51520206 |
| rs8034835 | Previous Literature | 15 | 51512664 |
| rs10046 | Previous Literature | 15 | 51502986 |
| rs2899472 | Previous Literature | 15 | 51516055 |
| rs1902586 | Previous Literature | 15 | 51570853 |

| | | | |
|---|---|---|---|
| rs3743205 | Previous Literature | 15 | 55790530 |
| rs77641439 | Previous Literature | 15 | 55722872 |
| rs12899331 | Previous Literature | 15 | 55801094 |
| rs1075938 | Previous Literature | 15 | 55790691 |
| rs57809907 | Previous Literature | 15 | 55722882 |
| rs6564903 | Previous Literature | 16 | 81653657 |
| rs11860694 | Previous Literature | 16 | 84457447 |
| rs1299348 | Previous Literature | 18 | 13822256 |
| rs11873029 | Previous Literature | 18 | 46617055 |
| rs8094327 | Previous Literature | 18 | 55963045 |
| rs12606138 | Previous Literature | 18 | 55993944 |
| rs459962 | Previous Literature | 21 | 15963120 |
| rs5965871 | Previous Literature | X | 144673082 |

The ALSPAC cohort genetic data used in this study was genotyped using the hg19 / GRHc 37 reference build. Position, thus, references the hg19 / GRHc 37 2014 build information for each SNP.

Supplemental Table S5

*Summary of regression tests of elastic net models for ALSPAC - Read*

| **Model** | **Adjusted R2** | **ANOVA for Model** | **ANOVA compared to null model** |
|---|---|---|---|
| SNP Only | 0.12 | F (53, 3593) = 10.32, p < .0001 | F (1, 53) = 10.32, *p* < .001 |
| SNP + Demographic | 0.32 | F (85, 3168) = 18.93, p < .0001 | F (1, 85) = 18.93, *p* < .001 |
